# Phylogenies increase power to detect highly transmissible viral genome variants

**DOI:** 10.1101/2023.07.28.23293332

**Authors:** Michael R May, Bruce Rannala

## Abstract

As demonstrated by the SARS-CoV-2 pandemic, the emergence of novel viral strains with increased transmission rates poses a significant threat to global health. Viral genome sequences, combined with statistical models of sequence evolution, may provide a critical tool for early detection of these strains. Using a novel statistical model that links transmission rates to the entire viral genome sequence, we study the power of phylogenetic methods—using a phylogenetic tree relating viral samples—and count-based methods—using case-counts of variants over time—to detect increased transmission rates, and to identify causative mutations. We find that phylogenies in particular can detect novel variants very soon after their origin, and may facilitate the development of early detection systems for outbreak surveillance.

---

The continuous emergence of novel genomic variants with the potential for increased transmissibility, virulence and other traits is a universal feature of viral pandemic and endemic diseases (*1*). The primary measure of transmissibility, *R*_0_, the basic reproductive rate, may be altered by many intrinsic and extrinsic factors (*2*). For example, D614G spike mutations in SARS-CoV-2 appear to enhance viral replication by increasing infectivity and stability of virions (*3*), epitope mutations may lead to immune escape increasing the population of susceptibles (*4*), mutations may cause increased viral loads in individuals due to enhanced replication increasing infectivity (*5*), or may cause a longer period of host infectivity (*6*). The theoretical models we explore in this study do not specify a mechanism for enhancing *R*_0_ but instead consider the most general case by assuming only that a change in *R*_0_ arises from a change in genome sequence.

Evidence for a causal role of one or more specific mutations in increasing the *R*_0_ of a strain (cluster of mutations) may come from a variety of sources, for example: epidemiological studies of case counts over time of individuals infected with particular strains (*7*); experimental studies of the transmissibility of different strains in an animal model system (*8, 9*); biophysical predictions – for example, predictions of binding affinities of mutant SARS-CoV-2 receptor binding domain (RBD) sequences to the ACE2 receptor based on molecular modeling (*10*); phylogenetic analysis of the expansions of particular strains through time (*11, 12*); etc. Although demonstrating enhanced transmission in an experimental animal system is often taken as the gold standard, no one source of evidence is definitive. Large differences in transmissibility may exist between human and model animal populations, casting doubt on the relevance of evidence from the model. On the other hand, epidemiological studies can directly infer enhanced transmission in human populations, but often sufficient data are available only after a variant has already become widespread (*13*).

The availability of genome sequences for infectious disease organisms, such as SARS-CoV-2, allows emerging mutations to be identified and monitored to assess their potential impacts (*14, 15*). Phylogenetic information defining evolutionary relationships among strains is also available from genome sequences and such data have been used with influenza, SARS-CoV-2, and other pathogens to predict the likely dominant variants (strains) of future pandemics (*16, 17*). Such methods implicitly assume that phylogenetic information provides additional predictive power beyond that available from simply monitoring changing frequencies of variants among infected individuals. However, surprisingly little is known about the relative power of phylogenies versus frequencies for identifying variants destined to become widespread.

Most genomic variants do not influence transmissibility and methods are urgently needed to identify (from among the hundreds or thousands of variants that do not influence transmissibility) the small subset of variants that do. Phylogenetic information from genome sequences could potentially be used for de novo identification of sites in viral genomes influencing transmissibility but theoretical studies are needed to understand the potential of such approaches. The statistical problem of identifying transmission enhancing sites is very similar to the challenging problem of identifying so-called “driver” mutations in genomes of cancer cells (*18*), although the smaller genomes of viruses greatly reduces the number of candidate mutations. Theoretical studies are needed both to demonstrate that such inferences are possible and to determine their prospective power.

Here we explore the information available from viral genomic datasets for early detection of transmission-enhancing mutations (referred to as TEs) that increase *R*_0_, as well as for identification of the specific sites with TE mutations in genomes. We focus on two types of data: 1) counts of viral variants sampled over time; 2) molecular phylogenies relating viral samples. To study information content, we develop an explicit statistical model of how mutation events influence transmissibility. Critically, this model allows us to derive tractable probability distributions for both count and phylogenetic datasets.

By simulating datasets based on realistic epidemiological and mutation parameters for SARS-CoV-2, we find that phylogenetic data provide strong evidence supporting TE status of variants days or weeks before case counts alone. This suggests that phylogenetic methods can identify emerging Variants of Concern (VOCs) or Variants of Interest (VOIs) sooner than methods using case counts. If epidemiological approaches for identifying VOCs based on case counts provide a lagging, or posthoc, indicator for the emergence of a new more transmissible variant, it is possible that phylogenetic analyses might allow candidate TE mutations to be identified before they are widespread enough for traditional epidemiological methods to estimate *R*_0_, thus providing a much-needed leading indicator for an emerging variant of interest.

## Modeling viral evolution with transmission-enhancing mutations

We present a theoretical exploration of the information content available from genomic sequence data under a simple branching process model that allows tractable calculations, and approximates (for large *R*_0_) the widely-used linear birth-death model of epidemic transmission dynamics (see Supplementary Material Section S1). We develop a basic model that allows one or more sites to potentially carry a TE nucleotide that influences the branching process underlying relationships of infecting lineages in a phylogenetic tree. Under this model, viral infections with genome sequence ***x*** infect new hosts (“transmit”) at rate *λ*(***x***), mutate to a new sequence with rate *ν* per site, and are sampled with rate *ϕ*. Viral lineages without any TE mutations have a base transmission rate of *λ*_0_; each TE mutation multiplicatively increases the transmission rate by a factor *δ*.

The posterior probability of model *M*, which specifies the specific base positions and nucleotides that confer a multiplicative increase of *R*_0_, can be explicitly calculated under our model using either complete phylogenetic information from the genome sequences, **X**, and sampling times, *T*, or instead using only counts of variant genomes (genotypes). The posterior probability obtained using phylogenetic information is

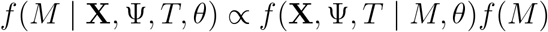

Here the phylogenetic tree and branch lengths, Ψ, are treated as known as well as the DNA mutation rates *ν*, baseline transmission rate *λ*_0_, and case sampling rate, *ϕ*, together represented as *θ* = {*ν, λ*_0_, *ϕ*}. We assume the TE effect size, *δ*, is unknown, and is integrated out in the above equation. The full details underlying this formula can be found in the Supplementary Material Section S2. It is important to note that we are calculating the joint probability of the data and phylogenetic tree where the shape of the phylogenetic tree is influenced by the TE site through its effect on the rate of transmission (birth events in the tree) but not by the other neutral sites which do not influence transmission rates. However, the probability distribution of the nucleotides observed at neutral sites does depend on the tree and branch lengths.

The probability of the model, *M*, when observing only genotype count data assumes that any phylogenetic information in the sampled genome sequences is ignored and these sequences are only used to obtain the counts of genotypes over time. While the phylogenetic tree is relevant to the count data because it determines how many copies of each genotype were present among all infected individuals when each of the samples were collected, it is not directly observed and we therefore treat it as an unobserved random variable. Thus, to obtain the marginal probability of the genotype counts, ***y*** = *g*(**X**), (which are a function of the sampled sequences as indicated) we integrate over the unobserved phylogenetic tree,

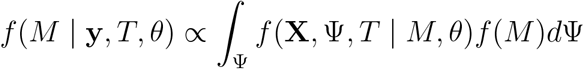

In practice, we use a direct numerical approach for evaluating the above equation rather than integration (see Supplementary Material Section S3).

## Phylogenetic data improve early detection of transmission-enhancing mutations

Early detection of variants with increased transmission rates is critical for mitigating outbreaks before they can become established. We used simulation to compare the ability of methods using either phylogenies or genotype counts to detect mutations that confer increased transmission in the early phase of an outbreak, assuming the TE site was known *a priori*. We simulated the first few weeks of outbreaks of pathogen variants bearing a single mutation that increased the transmission rate by a factor *δ* (the “effect size”). We varied the effect size over a range of plausible values of *R*_0_, from a 25% increase up to a doubling of *R*_0_ (see Supplementary Material Section S4). We then computed the posterior probability of a neutral model (where we assume that there was no TE site) and each possible TE model (corresponding to different TE nucleotides at the known TE site).

While both methods perform well for large effect sizes (Fig. 1, bottom row/left column, “both”), the tree method performs much better when the effect size is more modest (Fig. 1, left column, “tree only”); notably, the count method never succeeds when the tree method fails (Fig. 1, left column, “count only”). In cases when both methods succeed, the tree method detects increased *R*_0_ several days earlier than the count method (2.09 to 3.38 days on average for different effect sizes, *δ*) with detection using phylogenies occurring at least 9 days earlier in 5% of cases (with only a 25% increase of *R*_0_) (Fig. 1, middle column). The absolute time to detection depends on the effect size (Fig. 1, right column), ranging from over two weeks for modest effect sizes to about one week for the more extreme ones; as before, the tree method (orange lines) detects increased *R*_0_ earlier than the count method (blue lines). (We provide more details and results for this simulation in Supplemental Material Section S4).

**Figure 1:**
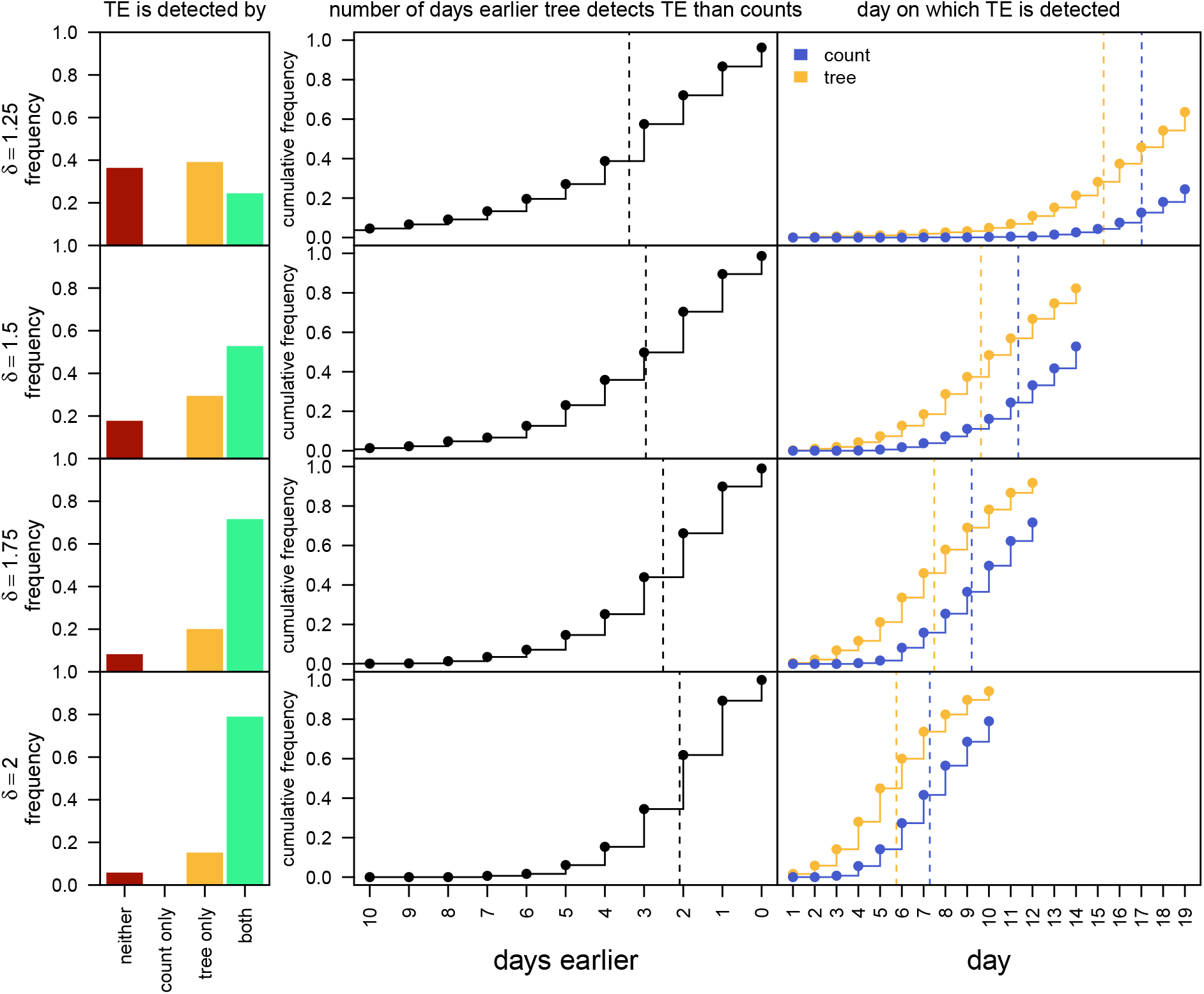
Phylogenies outperform counts at early detection. We simulated pathogen outbreaks where a single nucleotide change increased the transmission rate by a factor *δ* compared to the ancestral variant. We simulated each outbreak for a fixed number of days (10–19, depending on *δ*) after the origin of the novel variant, and compared the fit of a neutral model (the novel variant has the same transmission rate as the ancestral variant) against the true model (the given mutation confers an increased transmission rate) at daily increments. We measured the frequency with which the phylogenetic and count methods detected increased transmission (*i*.*e*., the true model had a posterior probability ≥95%) on at least one day of the outbreak (left column) as a function of the effect size (*δ*, rows). The count method never succeeds unless the tree method does as well (second bar), while the tree method often succeeds where the count method fails (third bar). If both methods succeeded, we computed: 1) the distribution (dots/lines) and mean (dashed lines) of the number of days earlier the tree method succeeded than the count method (center column), and; 2) and the distribution/mean for the day on which each method succeeded (right column). Overall, the phylogenetic method detects increased transmission several days earlier than the count method.

## De novo identification of transmission-enhancing mutations

In addition to quantitatively outperforming the count method when the TE variant is hypothesized *a priori*, the tree method also provides a practical approach for identifying *which* specific sites in the genome confer increased transmission rates (*i*.*e*., when TE sites are not know *a priori*). Indeed, the ability to scan a sample of sequenced viral genomes for evidence of variants with increased *R*_0_ is perhaps critical in identifying VOCs before other information about possible effects of variants (experimental studies, etc) are available.

We performed a simulation to characterize the ability of the tree method to correctly reject a neutral model (where no site in the genome confers increased transmission) and to identify the true site—and the specific nucleotide state that confers enhanced transmission—from among the entire genome. We simulated outbreaks with a single TE site over a range of effect sizes (as described previously) and with sample sizes ranging from 100 to 1600 viral samples. For each of these datasets, we also simulated the evolution of the neutral genome, comprising 29,999 sites (a genome size similar to SARS-CoV-2), using plausible values of the per-site mutation rate for SARS-CoV-2 (*19, 17*). We then computed the posterior probability of the neutral model and each single-site TE model (where a given TE model corresponds to a particular TE site/nucleotide combination).

Overall, the ability to decisively reject the neutral model increases as the effect size increases (Fig. 2, columns, blue lines) and as the number of samples increases (Fig. 2, *c, x*-axis). Beyond the ability to reject the neutral model, the tree method demonstrates generally good power to identify the true model. The frequency with which the true site/nucleotide combination has the highest support increases as a function of effect size, *δ* and sample size, *c* (Fig. 2, orange lines), exceeding 95% for TEs of large effect and large sample sizes; in many of these cases, the true model is decisively supported (*i*.*e*., it has a posterior probability greater than 95%; Fig. 2, red lines). (We provide more details and results for this simulation in Supplemental Material Section S4).

**Figure 2:**
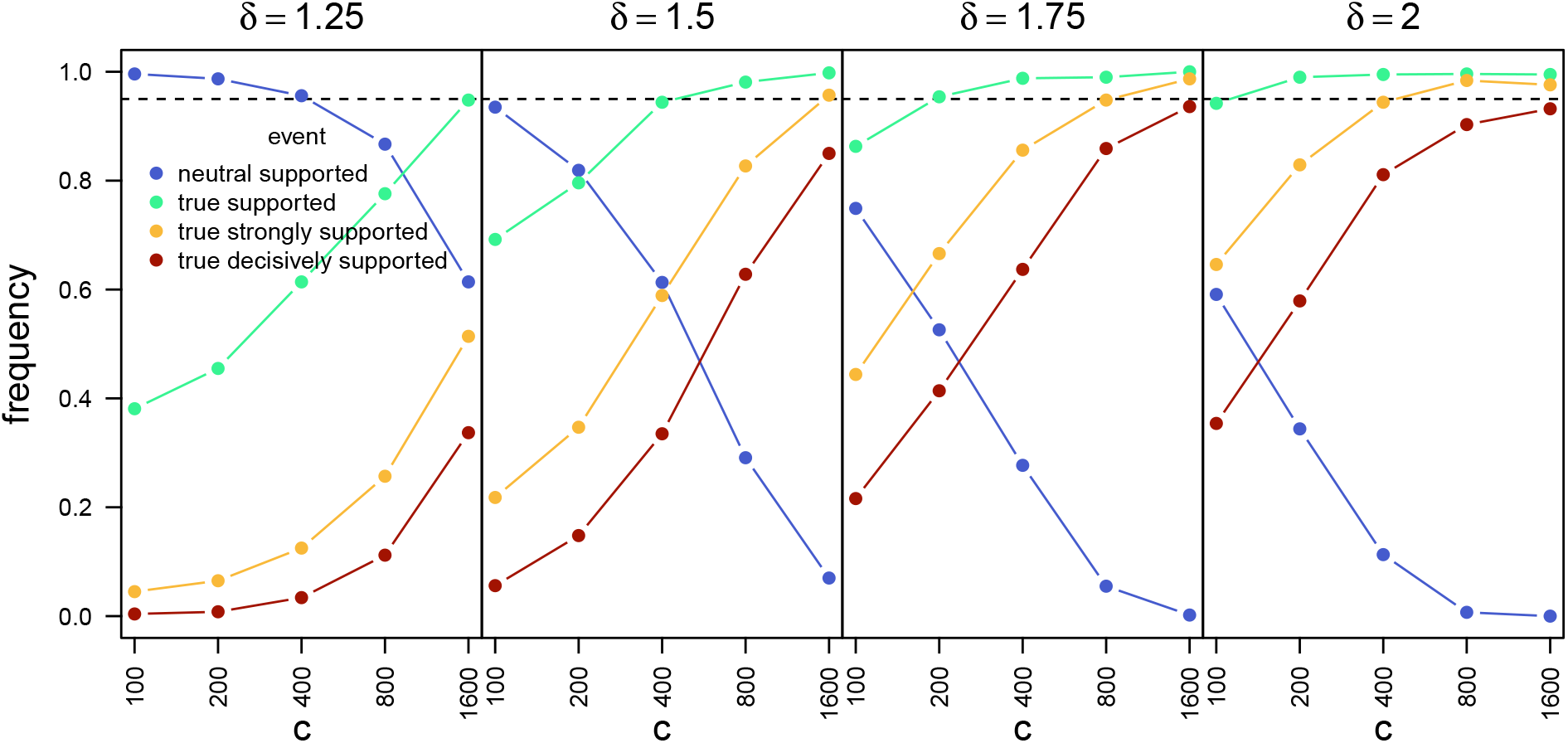
De novo identification of transmission-enhancing mutations. We simulated pathogen out-breaks in which a single nucleotide change increased the transmission rate by a factor *δ*. We then computed the support for a neutral model (where all variants have the same transmission rate) and all possible transmission-enhancing models (each corresponding to a particular genomic site/nucleotide combination). A model is supported if it has a posterior probability of at least 5%, strongly supported if it has a posterior probability higher than any other model, and decisively supported if it has a posterior probability greater than 95%. Across simulated datasets, the frequency with which the neutral model is supported (blue) decreases as a function of both the effect size (*δ*, columns) and the number of samples (*c, x*-axis). Conversely, the frequencies with which the true model is supported (green), strongly supported (yellow), or decisively supported increase as a function of the effect size and number of samples.

## Conclusions

We have developed a novel modeling framework to understand the theoretical behavior of methods for inferring changes in viral transmission rates caused by mutation events. The model incorporates several simplifying assumptions in order to make this study analytically and computationally tractable. For example, we have assumed that changes in transmission rates are driven by a single point mutation event; of course, multiple sites in the genome may confer increased transmission rates, and some transmission-enhancing variants may involve epistatic interactions among multiple sites, or recombination (*20, 21*). We have also assumed that patients do not recover (at least before the end of the monitoring date) and that viral sampling is stochastic and uniform; in reality, patients will recover over the course of days or weeks, and sampling effort naturally varies over time and space. While these assumptions may seem quite unrealistic, they may approximate the true process over short temporal and spatial scales, when multiple mutation and recombination events are unlikely, patient recovery is long relative to the monitoring period, and sampling effort is relatively homogeneous. In particular, these assumptions may be quite reasonable during the early stage of an epidemic or outbreak (*22*), which is the focus of our study. Nonetheless, state-dependent diversification models (*23*) could be developed that would provide the necessary mathematical and computational basis for incorporating more realistic models of mutation, recovery, and sampling; indeed, our theoretical exploration suggests that such efforts are worthwhile, as viral genomes appear to contain significant information about transmission-enhancing mutation events.

Our study demonstrates the theoretical and practical utility of phylogeny-based approaches for identifying emerging transmission-enhancing variants, which is a critical component of combating viral outbreaks. While experimental methods are currently the gold standard for identifying the mechanisms underlying changes in *R*_0_ caused by mutations (*9*), these approaches are expensive and time-consuming, and are difficult (if not impossible) to apply continuously and on short timescales. By contrast, mechanism-agnostic approaches—based on the frequency of genomic variants over time, or the phylogenetic relationship among variants— may provide earlier detection of variants with increased transmission rates before they become outbreaks. They may also identify specific variants that can then become targets of future experimental work.

We have demonstrated that, in theory, phylogeny-based approaches outperform frequency-based ones not only quantitively—they can detect an increased transmission rate for a given variant up to a week earlier, given realistic parameters for the SARS-CoV-2 pandemic—but also qualitatively: in contrast to frequency-based approaches, phylogenies allow us to detect increasing transmission rates when the variant is unknown *a priori*, even with relatively small sample sizes (on the order of hundreds or thousands of samples). These results support strategies for continuous sampling and genome sequencing of endemic viruses to monitor for emerging variants of concern (*24*). We are optimistic that theoretical and computational advances in phylogenetic approaches, and in particular stochastic modeling of transmission, mutation, and sampling processes, can lead to the development of early detection systems for monitoring and predicting epidemics that will be of significant value to the epidemiological community, and the world at large.

## Data Availability

This study does not involve any empirical data. All code used in this study is available at the at the GitHub release https://github.com/mikeryanmay/YuleSelectionArchive/releases/tag/initial_submission and the Zenodo DOI 10.5281/zenodo.8193217

## Acknowledgments

We are grateful to Brian R Moore, Jiansi Gao, Ammon Thompson, and X anonymous reviewers for their thoughtful discussion and feedback on this study.

## Authors contributions

M.R.M. and B.R. conceived the research, developed the statistical model, and wrote the manuscript. M.R.M. implemented the methods and conducted the simulation studies.

## List of Supplementary Materials

Supplemental Materials and Methods

Figs. S1 to S7

References (*24-41*)

