## Supplemental Materials and Methods for "Phylogenies increase power to detect highly transmissible viral genome variants"

#### S1 Stochastic Model of Virus Transmission Process

Our approach is based on the classical closed, spatially homogenous population SIR epidemic model proposed by (Kermack and McKendrick 1927). The population is composed of counts of individuals belonging to each of three classes: susceptibles $x$, infected (and circulating) $y$, and removed (recovered, dead or isolated) $z$. Using a deterministic (large population) model they derived the following differential equations of the process,

$$\begin{aligned}\frac{dx}{dt} &= -\beta xy,\\ \frac{dy}{dt} &= \beta xy - \gamma y,\\ \frac{dz}{dt} &= \gamma y.\end{aligned}$$

Kendall (1956) studied properties of the continuous time stochastic representation of this model with differential-difference equations,

$$p_{x,y,z}(t+\Delta t) = p_{x+1,y-1,z}(t)\beta(x+1)(y-1)\Delta t + p_{x,y+1,z-1}(t)\gamma(y+1)\Delta t + p_{x,y,z}(t)[1-(\beta x+\gamma)y\Delta t] + o(\Delta t)$$

Kendall (1956) noted that if the initial number of susceptibles, $m$, is large the population of infectious individuals can be studied in the early phase of the epidemic using a birth and death process with effective birth rate $\lambda = \beta m$ and death rate $\gamma$, simplifying the equations to be

$$p_y(t+\Delta t) = p_{y-1}(t)\lambda(y-1)\Delta t + p_{y+1}(t)\gamma(y+1)\Delta t + p_y(t)[1-(\lambda+\gamma)y\Delta t] + o(\Delta t).$$

Furthermore, as noted in Bailey, if we consider a sufficiently short time period relative to the recovery period and assume that quarantines are not in place the number of removals will be negligible and we can set $\gamma = 0$ approximating the epidemic dynamics of the emerging VOC as a Yule pure birth process with rate $\lambda$,

$$p_y(t) = p_{y-1}(t)\lambda\Delta t + p_y(t)[1-\lambda\Delta t] + o(\Delta t).$$

Our interest here is in the power of methods for early detection of VOCs so we will use a Yule process as an approximation of the epidemic transmission process throughout. Our Yule process model will be augmented, however, to include sampling of infected individuals and mutations among states of a transmission enhancing site of a DNA sequence through time.

Transmissibility of a pathogen is most often quantified using the basic reproductive rate, $R_0$ (Delamater et al. 2019). Dietz (1993) defines $R_0$ to be "the number of secondary cases one case would produce in a completely susceptible population." Since there is obviously stochastic variation of this number over individuals, others have defined $R_0$ as the average or expected number of secondary cases due to an infected individual (Diekmann et al. 1990). Using the expected number of new infections over the infective period of an individual as a definition, it is particularly simple to derive $R_0$ when using the birth-death approximation of Kendall (1956) because the birth and death rates are constants and thus $R_0$ is also. The goal is to find a mapping between the parameters of the linear birth-death process and the expected number of new infections per infected person, $R_0$. If a newly infected individual remains infective for time $t$ the conditional expectation of the number of birth events, $B$, (new infections caused by the individual) is

$$\mathbb{E}(B \mid t) = \lambda t. \tag{S.1}$$

The probability density of the time till recovery (or death) for a newly-infected individual is

$$f(t) = \gamma \mathrm{e}^{-\gamma t}. \tag{S.2}$$

The expected number of new infections caused by a newly-infected individual is then

$$R_0 = \int_0^\infty \lambda t \mathrm{e}^{-\gamma t} dt = \lambda \int_0^\infty t \mathrm{e}^{-\gamma t} dt = \frac{\lambda}{\gamma}. \tag{S.3}$$

In our analyses we are using a Yule process approximation that essentially assumes that $\gamma \to 0$ so the expectation is not defined. However, the expectation is essentially $\lambda \times (1/\gamma)$ where $\gamma$ will be a constant defined by the biological properties of the virus. Thus, if mutations do not influence the average duration of infectivity the relative birth rates for different variants will equal their relative $R_0$ values and meaningful comparisons therefore can be made using estimates of $\lambda$.

### S2 Model of Joint Sequence and Phylogeny Evolution

#### Defining the data and models

Consider a cumulative sample of $c$ genome sequences by time present, each of length $n$ with sites indexed $1, 2, \ldots, n$, and let $S$ be the set of site indexes for the genome. Let $N = \{\texttt{A}, \texttt{C}, \texttt{G}, \texttt{T}\}$ be the set of possible nucleotides at each site in the genome. Sequence sampling occurs over time and we let $\mathbf{X}(t) = \{x_{ij}(t)\}$ denote, at time $t$ in the past, the nucleotide at position $j$ of sequence $i$, where $x_{ij}(t) \in N$. We note that some lineage $i$ may not exist at a particular time $t$ (e.g., either $i$ was sampled prior to $t$ or was born after $t$) and in that case $x_{ij}(t) = \emptyset$ for all $j \in S$. We will also use the notation $\mathbf{X}$ for the sequence data and this represents a matrix of the state of each lineage at the time that it was sampled, this matrix has no empty elements. We consider a model in which nucleotides present at one or more sites in the genome may increase transmission rates for a pathogen, these are referred to as transmission-enhancing (TE) nucleotides; sites with one or more TE nucleotides are TE sites. We assume a particular site cannot be a TE site if it is not polymorphic; let $F \subset S$ be the set of indexes of polymorphic sites. The number of polymorphic sites is $d = |F|$, the cardinality of $F$. Since we will be interested only in polymorphic sites, we define the filtered dataset $\mathbf{X}^F = \{x_{ij}\}$ where $j \in F$. The set of indexes of invariant sites is the complement $I = F^C = G - F$. Denote the set of indexes for TE sites as $K \subset F$. The set of indexes of neutral (non-TE) polymorphic sites is the complement $W = K^C = F - K$. When there are no TE sites in the sampled genomes, $K = \emptyset$, otherwise, $K$ is a set of indexes, e.g., $K = \{1, 2, 3\}$ indicates that the first three sites are TEs. The cardinality $l = |K|$ is the number of TE sites. We define the filtered datasets $\mathbf{X}^I = \{x_{ij}\}$ where $j \in I$, $\mathbf{X}^K = \{x_{ij}\}$ where $j \in K$, and $\mathbf{X}^W = \{x_{ij}\}$ where $j \in W$. The sequence matrix with filter $J$ is defined at time $t$ to be $\mathbf{X}^J(t) = \{x_{ij}(t)\}$ where $j \in J$.

We consider a general multiplicative model where multiple alleles may be transmission enhancing at a given site. Denote the set of transmission-enhancing effect parameters for TE sites as $\Delta = \{\delta_j(x)\}$ where $j \in K$, $x \in N$, and $\delta_j(x) \in \mathbb{R}^+$. For each $j \in K$ we require $\delta_j(x) > 1$ for some $x \in N$ since otherwise the site is not a TE site. We also require $\delta_j(x) = 1$ for some $x \in N$ because at least one of the nucleotides must not influence birth rate. The number of parameters (dimension) of the TE model for the $j$th site depends on the number of nucleotides $x \in N$ for which $\delta_j(x) > 1$ and will vary from 1 to 3.

##### *The space of models*

A model is defined as a combination of the set of TE site indexes, $K$, and the nucleotide effects at those sites, $\Delta$,

$$M = \{K, \Delta\}.$$

The set of models is denoted $\mathcal{M} = \{M_i\}$.

The number of possible distinct sets of TE site indexes for $d$ polymorphic sites is the number of bipartitions of $d$, which is $2^d$. For a site in the set $K$, a model is uniquely defined by the set of nucleotides for which $\delta_j(x) > 1$. Under the conditions defined above, the possible number of TE nucleotides at any TE site is between 1 and 3, and the possible models for a TE site is then

$$p = 4 + \binom{4}{2} + \binom{4}{3} = 14.$$

The first term is the number of possible models with one TE nucleotide at the site, the second term is the number with two TE nucleotides, and so on. Given $j$ TE sites there are $\binom{d}{j}$ ways to assign the sites across the polymorphic sites and for each assignment there are $p^j$ possible models, therefore the total number of possible models with $d$ polymorphic sites is

$$|\mathcal{M}| = \sum_{j=0}^{d} \binom{d}{j} p^j = (1+p)^d = 15^d.$$

The number of possible models for $d$ polymorphic sites grows quickly with $d$. For example, with only $d = 10$ polymorphic sites there are $15^{10} \approx 5.8 \times 10^{11}$ distinct models. All these models are nested within the general model defined by $|K| = d$. The state space is greatly reduced by conditioning on the total number of TEs. With $|K| = 1$ there are only $pd + 1 = 14d + 1$ distinct models. For example, with $d = 10$ this corresponds to 141 distinct models. We can further reduce the state space of models by reducing the number of TE nucleotide models. Define the indicator function

$$\mathbb{I}[z] = \begin{cases} 1 & \text{if } z > 1 \\ 0 & \text{otherwise,} \end{cases}$$

and impose the constraint that for any site $j$,

$$\sum_{x_{\cdot j} \in N} \mathbb{I}[\delta_j(x_{\cdot j})] = 1,$$

so that each TE site has exactly one TE nucleotide. Then the number of possible models is $4d + 1$, which for $d = 10$ is only 41. We will explicitly consider results only under these simplified models in the analyses that follow.

#### Transmission rate of a sequence

Under a model with independent, multiplicative effects across sites, the birth (transmission) rate for the $i$th genome sequence $\mathbf{x}_i$ is

$$r(\mathbf{x}_i) = \lambda_0 \prod_{j \in K} \delta_j(x_{ij}),$$

where $\lambda_0$ is the birth (transmission) rate in the population ($R_0 - 1$ in time units of expected recoveries/deaths) for individuals infected with pathogen genomes containing no TEs. This is the model we assume in the analyses that follow, but it is easy to imagine other possible birth-rate functions, for example additive or subadditive models, or models that treat codons (rather than individual nucleotide sites) as transmission enhancing.

#### Stochastic process

We consider a viral transmission process that is described by a Yule process with rate $\Lambda = R_0 - 1$, where $R_0 > 1$, then $\Lambda \in (0, \infty)$. We imagine the process begins with a single viral lineage at time $t_0$ with sequence $\mathbf{X}(t_0) = \mathbf{x}_0(t_0)$. The process evolves forward in time under a Markov process with three types of events: mutation, transmission, and sampling.

Under this model, mutation events occur at rate $\nu$ per site per lineage, and are assumed to be independent across lineages and sites. When a mutation event occurs at a site, it mutates to each alternative nucleotide with equal probability, regardless of the current state. For the neutral sites, this is equivalent to the Jukes-Cantor substitution model, although a more general model (such as GTR) could be used instead. We assume that transmission events occur independently among lineages at a rate that depends on the current sequence of each lineage. For lineage $i$ with sequence $\mathbf{x}_i(t)$ at time $t$, the transmission rate is $r(\mathbf{x}_i(t))$. During a transmission event at time $t$, lineage $i$ splits into two daughter lineages, $j$ and $k$, each of which inherits the sequence $\mathbf{x}_i(t)$. We assume that lineages are sampled (and sequenced) with rate $\phi$ and that sampling removes the virus (infected individual) from the population so that no subsequent infections derive from that lineage. We allow this process to evolve forward until time $T$, at which point all remaining viral lineages are sampled and sequenced. This process gives rise to an unoberved phylogeny, $\Psi$, relating all of the $c$ samples, as well as the genome sequences of each sampled lineage, $\mathbf{X}$.

#### Phylogeny

The phylogeny, $\Psi = \{\mathbf{v}, B\}$, is a directed rooted binary tree with nodes (vertices) $\mathbf{v} = \{v_i\}$ and branches (edges) $b \in B$. Let $\mathbf{v}^A$ denote the set of ancestral nodes and $\mathbf{v}^T$ the set of tip nodes. For a phylogeny with $c$ samples, we index the tip nodes 1 through $c$, ordered according to the rows of the data matrix $\mathbf{X}$ so that $\mathbf{v}^T = \{v_i\}$ for $i \in \{1, 2, \ldots, c\}$. Starting with the root, we index the ancestral nodes $c+1$ through $2c-1$ so that $\mathbf{v}^A = \{v_i\}$ for $i \in \{c+1, c+2, \ldots, 2c-1\}$. A time is assigned to each node: the sampling time for tip nodes, and the inferred transmission time for ancestral nodes (measured forward in time). Let $\alpha(v)$ be the time of node $v$, $\alpha(\mathbf{v}^A)$ the times of the ancestral nodes, and $\alpha(\mathbf{v}^T)$ the times of the tip nodes. A branch $b \in B$ is a pair of nodes, $b = \{b_a, b_d\}$, with $b_a \in \mathbf{v}$ and $b_a \in \mathbf{v}$, ordered such that $\alpha(b_a) > \alpha(b_d)$, where we denote the ancestor node $b_a$ and the descendant node $b_d$. Because the branches specify ancestor-descendent relationship, they define the tree topology. The length of branch $b$ is:

$$t(b) = \alpha(b_d) - \alpha(b_a).$$

#### Likelihood

We are interested in calculating the joint probability of the sequences, $\mathbf{X}$, observed at all samples (present and past) and the phylogeny $\Psi$, as well as the TE model and the corresponding model parameters:

$$f(\mathbf{X}, \Psi \mid M, \lambda_0, \nu, \phi) = g(\mathbf{X}^K, \Psi \mid M, \lambda_0, \nu, \phi) h(\mathbf{X}^W, \mathbf{X}^I \mid \Psi, K, \nu), \tag{S.4}$$

where the first term on the right is the joint probability of the TE sites and the phylogeny, and the second term is the probability of the neutral sites (polymorphic and invariant), given the phylogeny. Note that $\mathbf{X} = \mathbf{X}^K \cup \mathbf{X}^W \cup \mathbf{X}^I$.

*Transition probabilities for TE sites.*—We represent the evolution of the TE sites along branches of the tree using a continuous-time Markov chain (CTMC). The joint probability of the TE sequences and the phylogeny can be obtained by calculating transition probabilities on branches and integrating over the unobserved TE sequences at internal nodes using a pruning algorithm (described below).

Because all demographic events (samplings or births) are observed for our Yule process model, to calculate the transition probabilities between sampling or birth events we only need to be able to calculate the probability that we start in state $\mathbf{x}^K(0)$ (corresponding to the sequence of TE sites at the beginning of a given branch $i$) and after time $t$ are in state $\mathbf{x}^K(t)$ (the sequence of TE sites at the end of branch $i$), with no birth or sampling events having occurred. There are $l = |K| + 1$ possible states for $\mathbf{x}^K(t)$. Namely, all possible sequences of length $|K|$ (the number of TE sites), plus an absorbing state $O$.

The instantaneous-rate matrix $\mathbf{Q} = \{q_{\mathbf{xy}}\}$ is an $l \times l$ matrix, whose element $q_{\mathbf{xy}}$ represents the instantaneous rate of change from filtered sequence $\mathbf{x}$ to filtered sequence $\mathbf{y}$. The instantaneous rate of change between two filtered sequences, $\mathbf{x}, \mathbf{y}$, is:

$$q_{\mathbf{xy}} = \begin{cases} \nu/3 & \text{if } \mathbf{x} \text{ and } \mathbf{y} \text{ differ by one nucleotide} \\ 0 & \text{otherwise} \end{cases}$$

and the rate of change between $\mathbf{x}$ and the absorbing state is:

$$q_{\mathbf{x}O} = r(\mathbf{x}) + \phi.$$

Finally, transitions out of the absorbing state have rate zero: $q_{O\mathbf{y}} = 0$ for all ending states $\mathbf{y}$. The transition probabilities can then be obtained by exponentiating the rate matrix:

$$p_{\mathbf{xy}}(t) = \left[ e^{\mathbf{Q}t} \right]_{\mathbf{xy}} D(\mathbf{y}),$$

where $\mathbf{x}$ and $\mathbf{y}$ are the filtered sequences at the beginning and end of the branch, $t$ is the branch length, and $D(\mathbf{y})$ is the probability density of the event at the end of the branch, which may depend on $\mathbf{y}$:

$$D(y) = \begin{cases} \phi & \text{if branch ends at sampling event} \\ 1 & \text{if branch ends at termination of process} \\ r(\mathbf{y}) & \text{if branch ends at internal node.} \end{cases}$$

To make explicit representation of the rate matrix straightforward, we simplify to a model of a single TE site with a single TE nucleotide, `A`, that enhances transmission by a factor $\delta$. In this case, $x^K(t) = \{\mathtt{A}, \mathtt{C}, \mathtt{G}, \mathtt{T}, O\}$, and the instantaneous-rate matrix $\mathbf{Q}$ is

$$\mathbf{Q} = \begin{pmatrix} - & \nu/3 & \nu/3 & \nu/3 & \delta\lambda_0 + \phi \\ \nu/3 & - & \nu/3 & \nu/3 & \lambda_0 + \phi \\ \nu/3 & \nu/3 & - & \nu/3 & \lambda_0 + \phi \\ \nu/3 & \nu/3 & \nu/3 & - & \lambda_0 + \phi \\ 0 & 0 & 0 & 0 & 0 \end{pmatrix},$$

with rows and columns corresponding to states of the state space and diagonal elements such that rows sum to 0. Note that transitions between pairs of nucleotides occur at equal rates. However, transitions to the absorbing state occur with rate $\delta\lambda_0 + \phi$ for `A` but with rate $\lambda_0 + \phi$ for all other nucleotides; transitions to the absorbing state occur faster when the lineage is in state `A` because transmission events occur at an elevated rate.

*Analytical transition probabilities for model with one TE site.*—Explicit solutions for the transition probabilities under the $\mathbf{Q}$ matrix defined above can be easily obtained via eigendecomposition. The eigenvalues are

$$\begin{aligned} E_1 &= 0 \\ E_2 &= -\lambda_0 - \phi - \frac{4}{3}\nu \\ E_3 &= -\lambda_0 - \phi - \frac{4}{3}\nu \\ E_4 &= -\frac{1}{6}(\beta + \alpha) \\ E_5 &= -\frac{1}{6}(\beta - \alpha) \end{aligned}$$

where we define

$$\begin{aligned}\alpha &= \sqrt{(\kappa+2\nu)^2 - 12(\delta-1)\lambda_0\nu},\\ \beta &= 3(\delta+1)\lambda_0 + 4\nu + 6\phi,\\ \kappa &= 3(\delta-1)\lambda_0 + 2\nu.\end{aligned}$$

and the matrix of eigenvectors is

$$\mathbf{U} = \begin{pmatrix} 1 & 0 & 0 & -\frac{(\alpha+\kappa)}{2\nu} & \frac{\alpha-\kappa}{2\nu} \\ 1 & -1 & -1 & 1 & 1 \\ 1 & 0 & 1 & 1 & 1 \\ 1 & 1 & 0 & 1 & 1 \\ 1 & 0 & 0 & 0 & 0 \end{pmatrix}$$

Now define the matrix

$$\mathbf{E} = \begin{pmatrix} 1 & 0 & 0 & 0 & 0 \\ 0 & e^{E_2 t} & 0 & 0 & 0 \\ 0 & 0 & e^{E_3 t} & 0 & 0 \\ 0 & 0 & 0 & e^{E_4 t} & 0 \\ 0 & 0 & 0 & 0 & e^{E_5 t} \end{pmatrix}$$

The matrix of transition probabilities is then obtained as

$$\mathbf{P}(t) = \mathbf{U}\mathbf{E}\mathbf{U}^{-1}.$$

The transition probabilities are given by the elements of $\mathbf{P}(t)$. If nucleotide A is the TE allele, we are interested in 4 distinct transition probabilities:

$$\begin{aligned}p_{AA}(t) &= \left(\frac{\alpha-\kappa}{2\alpha}\right)e^{-\frac{1}{6}t(\beta-\alpha)} + \left(\frac{\alpha+\kappa}{2\alpha}\right)e^{-\frac{1}{6}t(\beta+\alpha)},\\ p_{Ax}(t) &= p_{xA}(t) = \left(\frac{\nu}{\alpha}\right)e^{-\frac{1}{6}t(\beta+\alpha)}(e^{\frac{1}{3}t\alpha}-1),\\ p_{xx}(t) &= \left(\frac{\alpha-\kappa}{6\alpha}\right)e^{-\frac{1}{6}t(\alpha+\beta)} + \left(\frac{\alpha+\kappa}{6\alpha}\right)e^{-\frac{1}{6}(\alpha-\beta)} + \frac{2}{3}e^{-t(\lambda_0+\phi+\frac{4}{3}\nu)},\\ p_{xy}(t) &= \left(\frac{\alpha-\kappa}{6\alpha}\right)e^{-\frac{1}{6}t(\alpha+\beta)} + \left(\frac{\alpha+\kappa}{6\alpha}\right)e^{-\frac{1}{6}t(\alpha-\beta)} - \frac{1}{3}e^{-t(\lambda_0+\phi+\frac{4}{3}\nu)}.\end{aligned}$$

*Likelihood of TE sites and phylogeny.*—Having defined transition probabilities for TE sites, we now turn to computing the joint likelihood of a filtered sample, $\mathbf{X}^K$, and the rooted phylogeny, $\Psi$, $g(\mathbf{X}^K, \Psi \mid M, \lambda_0, \nu, \phi)$. We can compute the joint probability of the sequences and phylogeny if we assume we know the sequences of the ancestral nodes in the same order that they appear in $\mathbf{v}^A$). The joint probability of the sequence and the phylogeny is the probability of a given ancestral-state configuration, summed over all such configurations:

$$g(\mathbf{X}^K, \Psi \mid M, \lambda_0, \nu, \phi) = \sum_{\mathbf{x}_1^K(\alpha[1])} \sum_{\mathbf{x}_2^K(\alpha[2])} \cdots \sum_{\mathbf{x}_{c-1}^K(\alpha[2])} \pi[\mathbf{x}_1^K(\alpha[1])] \prod_{b\in B} p_{\mathbf{x}_{b_a}^K(\alpha[b_a]) \to \mathbf{x}_{b_d}^K(\alpha[b_d])}\big(t(b)\big),$$

where $\pi[\mathbf{x}_1^K(\alpha[1])]$ is the probability that the root is in state $\mathbf{x}_1^K(\alpha[1])$, and $p_{\mathbf{x}_{b_a}^K(\alpha[b_a]) \to \mathbf{x}_{b_d}^K(\alpha[b_d])}\big(t(b)\big)$ is the transition probability from the ancestral to descendant sequence along branch $b$. To be concise, we suppress the dependence of $p_{i\to j}(t)$ on $M$, $\lambda_0$, $\nu$ and $\phi$ in our notation. For simplicity we assume that all root states are equally likely. This sum can be efficiently evaluated using Felsenstein's pruning algorithm Felsenstein (1981).

*Likelihood of neutral sites.*—The likelihood of the neutral sites under the Jukes-Cantor model, $h(\mathbf{X}^W, \mathbf{X}^I \mid \Psi, \nu)$, can be calculated using standard phylogenetic methods (Felsenstein's pruning algorithm; Felsenstein 1981).

#### Likelihood under a purely neutral model

In the simulation study that follows, we investigate the ability of the phylogenetic model to correctly identify the TE model. To do so, we compare among many candidate TE models, as well as a model where there is no TE site (a purely neutral model). The likelihood of the data under this model is:

$$f(\mathbf{X}, \Psi \mid \lambda_0, \nu, \phi) = q(\Psi \mid \lambda_0, \nu, \phi) h(\mathbf{X}^W, \mathbf{X}^I \mid \Psi, K, \nu), \tag{S.5}$$

where $q(\Psi \mid \lambda_0, \nu, \phi)$ is the probability of the tree under a Yule model with sampling events.

To compute the likelihood under this model, we specify a 2-state Markov model, where the first state corresponds to the process having a single, unsampled lineage, and the second state is an absorbing state. The corresponding rate matrix matrix $\mathbf{R}$ is:

$$\mathbf{R} = \begin{pmatrix} -(\lambda_0 + \phi) & \lambda_0 + \phi \\ 0 & 0 \end{pmatrix}$$

The probability of starting with one lineage and ending with one lineage over a duration of length $t$ is $p_{11}(t) = \left[ \mathrm{e}^{\mathbf{Q}t} \right]_{11}$, where the subscript $_{11}$ the element of the first row and first column of the matrix $\mathrm{e}^{\mathbf{Q}t}$. The transition probability along a branch in a tree is:

$$p_{11}(t) = d\mathrm{e}^{-(\lambda_0 + \phi)t},$$

where the quantity $d$ is a point density for the event at the end of the branch:

$$d = \begin{cases} \phi & \text{if branch ends at sampling event} \\ \lambda_0 & \text{if branch ends at internal node} \\ 1 & \text{if branch ends at termination of process (no event).} \end{cases}$$

The probability of the tree is the product of such transition probabilities across branches:

$$q(\Psi \mid \lambda_0, \nu, \phi) = \prod_{b \in B} p_{11}\big(t(b)\big).$$

### S3 Model of Count Evolution

The stochastic model described in Section S2 provides a complete description of the probability model underlying the complete data structure $\mathbf{X}(t)$ which defines the state of the sequence of each lineage existing at time $t$ as well as which lineages are born, or are sampled at that instant in time. We consider two likelihoods: the likelihood of the sampled sequences conditional on a phylogenetic tree with branch lengths (that specify the sequence of birth and sampling events), and; the likelihood of the sequences (genotypes) sampled at each sampling event. Both of these likelihoods are obtained by marginalizing over some aspects of the complete data $\mathbf{X}(t)$. In the case of the phylogenetic model, we assume we know the tree but not the states of the sequences other than at the tips of the tree (the sampling times). Thus, the phylogenetic likelihood is obtained by marginalizing over the sequence states along the branches and at the vertices of the phylogeny; this is done by calculating transition probabilities between states at vertices and summing (marginalizing) over all possible nucleotide states at each internal vertex of the tree. Note that the phylogenetic tree specifies the number of lineages that exist at any point in time (including the sampling times) and the genealogical relationships among lineages but does does not specify the sequence type of each lineage except at the sampling times (see the left panel of Figure S1). Sequence types are indicated by colors in the figure.

The count data does not include information about the number of sequences that exist at any time; in particular, at each sampling time all that is known is that at least two lineages existed at sampling time $t$ if this is not the last sampling event (including the sampled lineage). Also, when a lineage is sampled, its relationship to other sampled lineages is unknown. Thus, the likelihood of the count data is obtained by marginalizing over the phylogeny as well as the sequence states on lineages:

$$f(\mathbf{X}, \alpha(\mathbf{v}^T) \mid M, \lambda_0, \nu, \phi) = \sum_{B \in \mathcal{B}} \int_{\alpha(\mathbf{v}^A) \in \mathcal{T}_B} f(\mathbf{X}, \Psi = \{\mathbf{v}, B\} \mid M, \lambda_0, \nu, \phi) \, \mathrm{d}\alpha(\mathbf{v}^A), \tag{S.6}$$

where $\mathcal{B}$ is the set of all possible ancestor-descendant relationships (topologies) and $\mathcal{T}_B$ is the set of permissible ancestral node ages for a given topology, constructed as

$$\mathcal{T}_B = \left\{ \alpha(\mathbf{v}^A) \; : \; t(b) \in \mathbb{R}^+ \; \forall \, b \in B \right\},$$

that is, we require that all branch lengths are positive. The integrand in equation (S.6) is the joint probability of the phylogeny and sequences, equation (S.4), which integrates over the sequence states of ancestral lineages.

In practice, it is easier to consider an intermediate marginalization, specifically of the set of lineages existing at the time of each sampling event and their genotype (sequence state) (Figure S1, middle panel). This is achieved by imagining the unsampled lineages and their genotypes at each sampling time are known (Figure S1, right panel). We can then sum over all possible configurations of unsampled lineages (which are treated as latent variables and subsequently referred to as latent lineages). The result is the marginal probability of the counts. The remaining sections focus on the details of these calculations, which are solved numerically.

##### Model of Genotype Count Evolution

We develop a birth-mutation-sampling process that models counts of different genotypes over time (*i.e.*, the number of samples of each genotype over time). This process has the same parameters as those of the phylogenetic model described above and has the same diversification and mutation dynamics; rather than changing the underlying evolutionary process, we are just observing different

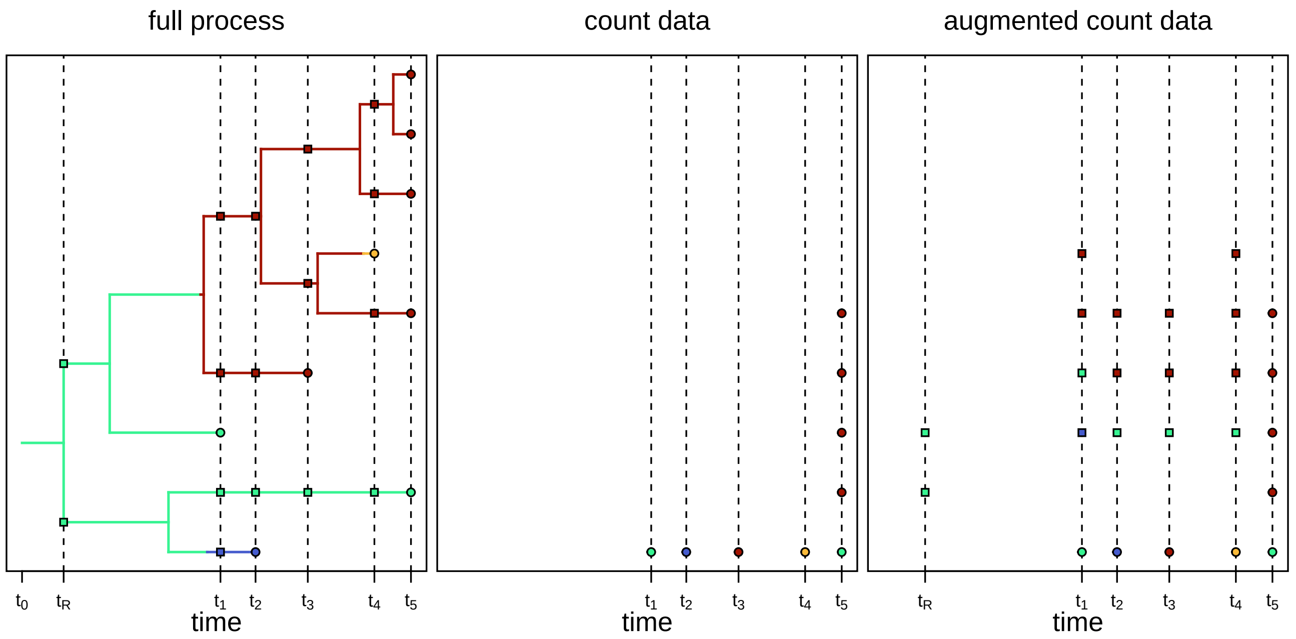


**Figure S1: Relationship between phylogenetic data and count data.** Left) The birth-mutation-sampling process produces a phylogenetic tree (left) with branches colored by the genotype at each time (the "full process"). This process produces samples of particular genotypes (circles at tips, colored according to genotype) at particular times, $t_i$ ($x$-axis, and indicated by vertical dashed lines). The time $t_R$ is the time of the first birth event rather than the time of a sample. Middle) The phylogenetic data can be translated into count data by counting the number of samples of each genotype at each sample time (*i.e.*, at each vertical dashed line). Right) We augment the count data with "latent" (unobserved) lineages (squares, colored according to genotype), which represent the unknown number of unsampled lineages at each sample time. Given the unknown phylogeny (left), the latent counts at time $t_i$ can be determined by the color of each branch that intersects the corresponding vertical line. Note that we also include the number of lineages at the first bifurcating event, $t_R$, in the augmented data.

variables. Our approach to modeling count data is very similar to previous work on birth-death-migration processes (Bailey 1968; Renshaw 1972), where mutation events correspond to migration events between genotype-specific subpopulations. While birth-death-migration processes are often quite intractable, our birth-mutation-sampling model allows a number of simplifications that render it amenable to numerical analysis. We begin by presenting the full model of genotype subpopulations, and then work through simplifications that lead to tractable, exact results for a model with a single bi-allelic TE site.

##### *Stochastic process*

We model the evolution of a population of lineages defined by counts of distinct genotypes. Let $G_i$ denote the distinct viral sequence that is genotype $i$, and $\mathcal{G}$ the set of all possible unique genotypes. There are $g = |\mathcal{G}| = 4^n$ such distinct genotypes which define genotype-specific subpopulations. The subpopulations evolve under a birth and sampling process with genotype-specific birth rates, and also exchange "migrants" during mutation events (a mutation event in our model corresponds to a lineage migrating from one subpopulation to another in the classical multitype migration models).

We number the distinct genotypes arbitrarily from 1 to $g$. We denote the count of individuals infected with viral genotype $i$ at time $t$ as $y_i(t)$. The state of the process at time $t$ is thus a $g$-tuple of counts, $\boldsymbol{y}(t) = \{y_i(t)\}$ for all $i \in \mathcal{G}$. The process begins at time $t_0$ with a single lineage of genotype $i$, namely $y_i(t_0) = 1$ and $y_{j \neq i}(t_0) = 0$, and evolves forward in time until some pre-specified end time (the final sampling event), $t_e > t_0$. Genotype counts evolve under a birth-mutation-sampling process. The total rate at which lineages of genotype $i$ give birth is $r(G_i)y_i(t)$, where $r(G_i)$ is the birth rate for a sequence matching genotype $G_i$. During a birth event, one lineage of genotype $i$ gives birth to a new lineage, increasing $y_i(t)$ by one. Sampling events occur at rate $\phi y_i(t)$. During a sampling event, a lineage of genotype $i$ dies, $y_i(t)$ decreases by one, and the time and genotype state are recorded. Finally, mutation events away from genotype $i$ occur at rate $\nu y_i(t)$. During a mutation event, a lineage of genotype $i$ has a site within its genome is chosen uniformly at random; the chosen nucleotide is then mutated to one of the three other nucleotides chosen uniformly at random. The mutation event causes the lineage, currently with genotype $G_i$, to mutate to a new genotype, $G_j$. As a result, $y_i(t)$ decreases by one and $y_j(t)$ increases by one. We stop the process at time $t_e$, and sample all the lineages alive at that point, recording their sampling times ($t_e$) and genotype states.

The outcomes of this process are the time at which each sample is taken, $\alpha(\mathbf{v}^T)$ in our phylogenetic notation (or the set $T$ in our count notation), and the sequences (genotypes) belonging to each sample, namely $\mathbf{X}$. The genotype counts are a function of the sampled sequences, $\mathbf{X}(t)$ (defined earlier), namely

$$y_i(t) = \sum_{j=1}^{c} \mathbb{I}_i[\mathbf{x}_j(t)],$$

where

$$\mathbb{I}_i[\mathbf{x}_j(t)] = \begin{cases} 1 & \text{if } \mathbf{x}_j(t) = G_i \\ 0 & \text{otherwise} \end{cases}$$

If $t$ is a sampling time, the genotype sampled at time $t$ is defined by vector subtraction as

$$z(t) = \boldsymbol{y}(t) - \boldsymbol{y}(t - \Delta t),$$

where $\Delta t$ is sufficiently small that a single lineage is sampled during time interval $(t - \Delta t, t)$ with probability arbitrarily close to 1. The result is a vector with a single non-zero element. Since indexes correspond to genotype labels this index is retained as the variable $z(t)$ which uniquely identifies the genotype of the lineage sampled at time $t$. The final sample at time $t_e$ is the complete configuartion of genotypes existing at time $t_e$, namely $\boldsymbol{y}(t_e)$. Define the sample data (excluding the last sample) to be $\boldsymbol{z} = \{z(t)\}$ for $t \in T$ and $t \neq t_e$. The complete data are then $T$, $\boldsymbol{z}$ and $\boldsymbol{y}(t_e)$.

#### Likelihood of genotype count data

Our goal is to compute the likelihood of the observed genotype count data, $f(\boldsymbol{z}, \boldsymbol{y}(t_e), T \mid M, \lambda_0, \nu, \phi)$. In contrast to the stochastic process described above, in real use cases the origin time of the process, $t_0$, is unknown *a priori*. We therefore include an additional parameter, $t_R$, which represents the time of the initial bifurcating event (the root of the phylogeny), and compute the conditional likelihood, $f(\boldsymbol{z}, \boldsymbol{y}(t_e), T \mid M, \lambda_0, \nu, \phi, t_R)$; we use the root time rather than the origin time to maintain equivalence with the phylogenetic likelihood. We integrate out the root time, which is unknown based on count data, using a numerical procedure described in a subsequent section.

##### *Count data structures*

The sample times, $\alpha(\mathbf{v}^T)$ and sequence data, $\mathbf{X}$, need to be mapped into genotype count data, $\mathbf{z}$ and $T$ (see Figure S1). First, $\alpha(\mathbf{v}^T)$ includes redundant values because samples taken at the end of the process all have identical ages. We denote the ordered set of distinct sample times as $T = \{t_i = \alpha(v_i)\}$ for all $i \in \{1, \ldots, c\}$ (note that placing the times in a set removes duplicates), with element $t_i$ being the $i^{\text{th}}$ distinct sample time. The set is arranged in ascending order such that $t_{i-1} < t_i$. We denote the number of distinct samples times $s = |T|$.

The sampled genotypes $\boldsymbol{z}$ are indexes in the set $\{1, \ldots, g\}$ and the latent lineages $\boldsymbol{y}(t)$ are whole numbers, $\mathbb{W}$ (counts of genotypes), and make up elements of the space $\mathbb{W}^g$. This number of elements in this space equals the number of genotypes and each element takes values on a countably infinite set since the number of lineages with genotype $i$ can grow to infinity. However, for any given realization, we know that there could not have been more than $c$ lineages over the entire history of the realization because all lineages are ultimately sampled (either stochastically or at the end of the process). For a given realization with $c$ sampled genomes, we can thus construct a finite subspace $\Omega^g \subset \mathbb{W}^g$ where $\Omega = \{0, 1, \ldots, c\}$ such that $\sum_i^g \boldsymbol{y}_i(t) \leq c$ for all count vectors $\omega^g \in \Omega^g$. For a given number of lineages $k$, there are $\binom{k+g-1}{g-1}$ ways to distribute those lineages among the $g$ genotypes. Therefore, the number of distinct sample configurations in this subspace (its cardinality) is:

$$|\Omega^g| = \sum_{k=0}^{c} \binom{k+g-1}{g-1} \tag{S.7}$$

*Augmented likelihood of counts*

We now consider the joint probability of the sampled genotypes $\boldsymbol{z}$ and latent lineages $\boldsymbol{y}(t)$ observed at times $T$, expanding the state space to include (unobserved) genotype counts of unsampled (latent) lineages. We use the notation $\boldsymbol{y}_T = \boldsymbol{y}(t)\ \forall t \in T$ to represent the latent counts at the sampling times. We consider the probability of the genotype, $z(t_i)$, sampled at time $t_i$ given the state of latent lineages at the previous sampling time $t_{i-1}$,

$$\begin{aligned} p(z(t_i), \boldsymbol{y}(t_i) \mid \boldsymbol{y}(t_{i-1})) &= \lim_{\Delta t \to 0} p(z(t_i) \mid \boldsymbol{y}(t_i - \Delta t)) p(\boldsymbol{y}(t_i - \Delta t) \mid \boldsymbol{y}(t_{i-1} + \Delta t)) \\ &= p(z(t_i) \mid \boldsymbol{y}_-(t_i)) p(\boldsymbol{y}_-(t_i) \mid \boldsymbol{y}_+(t_{i-1})) \end{aligned}$$

where the $+$ and $-$ subscripts indicate the limits on the left and right, respectively. The probability density is not continous at the sampling event and therefore the two limits are not equal. The joint density of the latent counts and sampled genotypes is

$$\begin{aligned} f(\boldsymbol{z}, \boldsymbol{y}(t_e), \boldsymbol{y}(t_R), \boldsymbol{y}_T, T \mid M, \lambda_0, \nu, \phi, t_R) = \pi\big(\boldsymbol{y}(t_R)\big) \prod_{i=2}^{s-1} p(z(t_i) \mid \boldsymbol{y}_-(t_i)) p(\boldsymbol{y}_-(t_i) \mid \boldsymbol{y}_+(t_{i-1})) \\ \times\, p(\boldsymbol{y}(t_e) \mid \boldsymbol{y}_+(t_{s-1})) p(z(t_1) \mid \boldsymbol{y}_-(t_1)) p(\boldsymbol{y}_-(t_1) \mid \boldsymbol{y}(t_R)). \end{aligned}$$

This probability is depicted in Fig. S2.

*Likelihood of counts*

We marginalize the joint probability over all possible values of $\mathbf{y}(t)\ \forall t \in T$ and the root lineages to yield the likelihood of the counts:

$$f(\boldsymbol{z}, \boldsymbol{y}(t_e), T \mid M, \lambda_0, \nu, \phi, t_R) = \sum_{\boldsymbol{y}(t_R)} \sum_{\boldsymbol{y}_T} f(\boldsymbol{z}, \boldsymbol{y}(t_e), \boldsymbol{y}(t_R), \boldsymbol{y}_T, T \mid M, \lambda_0, \nu, \phi, t_R)$$

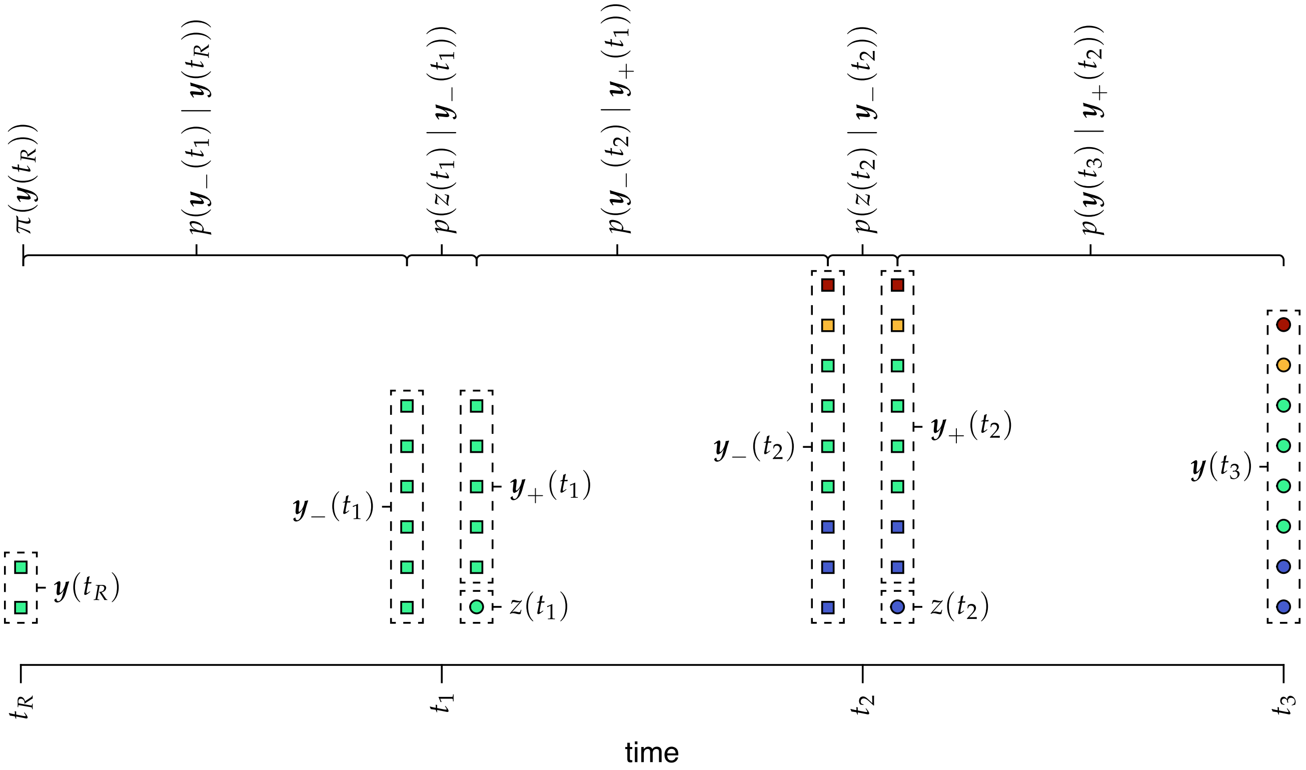


**Figure S2: Likelihood of the augmented count data.** To compute the augmented likelihood of the count data, we imagine the number of lineages at each time point, $\boldsymbol{y}(t)$, is known (*i.e.*, the count dataset is augmented with the number of lineages). The process begins with lineages $\boldsymbol{y}(t_R)$ with probability $\pi(\boldsymbol{y}(t_R))$. These lineages diversify over the first time interval (up to moment $t_1$) to produce the lineages $\boldsymbol{y}_-(t_1)$ some infinitesimal amount of time before the first sampling event. The lineages $\boldsymbol{y}_-(t_1)$ then produce a sample at time $t_1$; this removes the chosen sample $z(t_1)$ from the process and leaves $\boldsymbol{y}_+(t_1)$ to diversify into the next time interval. This process continues forward in time until the last time point, $t_3$, where all lineages are collected. The augmented likelihood is the product of transition probabilites between time intervals and the probability densities of the sampling events.

where the sums are over all possible values of $\boldsymbol{y}(t_R)$ and $\boldsymbol{y}_T$ the joint set of latent counts at each sampling event except the last. The sums and products in this equation can be re-arranged as:

$$f(\boldsymbol{z}, \boldsymbol{y}(t_e), T \mid M, \lambda_0, \nu, \phi, t_R) = \sum_{\boldsymbol{y}(t_R)} \pi\big(\boldsymbol{y}(t_R)\big) \sum_{\boldsymbol{y}(t_1)} p(\boldsymbol{y}_-(t_1) \mid \boldsymbol{y}(t_R)) p(z(t_1) \mid \boldsymbol{y}_-(t_1))$$
$$\times \sum_{\boldsymbol{y}_T} \left[ \prod_{i=2}^{s-1} p(\boldsymbol{y}_-(t_i) \mid \boldsymbol{y}_+(t_{i-1})) p(z(t_i) \mid \boldsymbol{y}_-(t_i)) \right] p(\boldsymbol{y}(t_e) \mid \boldsymbol{y}_+(t_{s-1})).$$

Here the sums are over the left or right limit as defined in the terms of the equations.

To evaluate this equation, we introduce the conditional likelihood vector $l\big(\boldsymbol{y}(t)\big)$, which represents the probability of the younger data, conditional on the process currently being in state $\boldsymbol{y}(t)$. Again, we use $l\Big(\boldsymbol{y}_+(t)\Big)$ and $l\Big(\boldsymbol{y}_-(t)\Big)$ to represent the conditional likelihood given the process is in state $\boldsymbol{y}(t)$ immediately before or after a sampling event, respectively.

We begin by filling in the conditional likelihood vector at the second-to-last sample time, $t_{s-1}$:

$$l\Big(\boldsymbol{y}_-(t_{s-1})\Big) = p\Big(\boldsymbol{y}(t_s) \mid \boldsymbol{y}_-(t_{s-1})\Big)$$

for each possible value of the latent lineages, $\boldsymbol{y}_-(t_s)$. This is the probability of transition from state $\boldsymbol{y}_-(t_{s-1})$ immediately after the last sampling event to the lineages sampled at the present.

We then step back to the moment immediately before the sampling event and compute:

$$l\Big(\boldsymbol{y}_+(t_{s-1})\Big) = \sum_{\boldsymbol{y}_-(t_{s-1})} p\Big(\boldsymbol{y}_-(t_{s-1}) \mid \boldsymbol{y}_+(t_{s-1})\Big),$$

again for each possible value of the latent lineages.

We then recursively step back to each prior sample time point $i$, repeating these two steps. First, we compute the transition probability:

$$l\Big(\boldsymbol{y}_-(t_i)\Big) = \sum_{\boldsymbol{y}_+(t_{i-1})} p\Big(\boldsymbol{y}_+(t_{i-1}) \mid \boldsymbol{y}_-(t_i)\Big).$$

Next, we compute the sampling probability density:

$$l\Big(\boldsymbol{y}_+(t_i)\Big) = \sum_{\boldsymbol{y}_-(t_i)} p\Big(\boldsymbol{y}_-(t_i) \mid \boldsymbol{y}_+(t_i)\Big).$$

Because we enumerate each possible value of future latent lineages at each step, we are averaging over all possible values of the latent lineages as we move backward toward the root.

We repeat this recursion up to (and including) the root at time $t_R$. The unconditional likelihood is then:

$$f(\boldsymbol{z}, \boldsymbol{y}(t_e), T \mid M, \lambda_0, \nu, \phi, t_R) = \sum_{\boldsymbol{y}(t_R)} \pi\big(\boldsymbol{y}(t_R)\big) l\big(\boldsymbol{y}(t_R)\big), \tag{S.8}$$

*i.e.*, we compute the likelihood of the data given each possible root state, and average over the root states in proportion to their initial probability.

*Matrix notation.*—We can represent the unconditional likelihood more compactly using matrix notation. In this case, we represent the initial probability at the root as a column vector, $\boldsymbol{\pi}$, which is a vector of probabilities, one per root state. We represent the transition probabilities between sampling points as transition probability matrices, $P(\tau) = \{p_{\boldsymbol{x},\boldsymbol{y}}\}$, where the element $p_{\boldsymbol{x},\boldsymbol{y}}(\tau)$ is the probability of transitioning from lineages $\boldsymbol{x} \in \Omega^g$ to $\boldsymbol{y} \in \Omega^g$ over an interval of length $\tau$. Finally, we represent the sampling densities as matrices $S(t) = \{s_{\boldsymbol{xy}}\}$, where element $s_{\boldsymbol{xy}}(t)$ is the probability density of the sampling event that begins with lineages $\boldsymbol{x} \in \Omega^g$ and ends with lineages $\boldsymbol{y} \in \Omega^g$ (and therefore produces observed counts $\boldsymbol{x} - \boldsymbol{y}$).

The matrix representation of the unconditional likelihood is then

$$f(\boldsymbol{z}, \boldsymbol{y}(t_e), T \mid M, \lambda_0, \nu, \phi, t_R) = \boldsymbol{\pi} P(t_1 - t_R) S(t_1) \left[\prod_{i=2}^{s-1} P(t_i - t_{i-1}) S(t_i)\right] P(t_s - t_{s-1}) I, \tag{S.9}$$

where $I$ is a row vector with values

$$I(\boldsymbol{\omega}^g) = \begin{cases} 1 & \text{if } \boldsymbol{\omega}^g = z(s) \\ 0 & \text{otherwise} \end{cases},$$

*i.e.*, it is a vector that zeros out transitions to states at the end of the process that do not match the observed counts at that time.

*Initial probability.*—The initial probability represents the probability density that the process began with a single lineages of a given genotype, which then experienced a birth event. In principle, this can be any function that assigns positive probability density only to initial lineage vectors with exactly two lineages of the same genotype (because a birth event just occurred to a single lineage in the given genotype, giving rise to two lineages in the same genotype). In practice, we will assume the first bifurcating event occurred to a lineage in a particular genotype, $G$; *i.e.*, we will imagine that the investigator has a good idea about the ancestral genotype. In this case, the initial probability function is

$$\pi(\boldsymbol{y}(t_R)) = \begin{cases} r(G) & \text{if } y_G(t_R) = 2, \text{ and } y_H(t_R) = 0 \text{ for all } H \in \mathcal{G} \neq G \\ 0 & \text{otherwise.} \end{cases}$$

Note that the probability density associated with this state is $r(G)$ (the birth-rate function evaluated for genotype $G$), because this is the probability density of a birth event for the ancestral lineage.

*Transition probabilities.*—The transition probability, $p\big(\boldsymbol{y}(t_2) \mid \boldsymbol{y}(t_1)\big)$, is the probability of transitioning from lineages $\boldsymbol{y}(t_1)$ to lineages $\boldsymbol{y}(t_2)$ over a time interval of duration $(t_2 - t_1)$ without leaving any samples.

Before we proceed, it will be convenient to introduce some functions for rates of events for the entire population (rather than per individual or per genotype). The total birth rate for a given population is

$$\Lambda(\boldsymbol{\omega}^g) = \sum_{i=1}^{g} r(G_i)\boldsymbol{\omega}_i^g,$$

*i.e.*, it is the sum of the birth rates across individuals in the entire population. Likewise, the total sampling rate is

$$\Phi(\boldsymbol{\omega}^g) = \sum_{i=1}^{g} \phi \boldsymbol{\omega}_i^g(t).$$

and the total mutation rate is

$$N(\boldsymbol{\omega}^g) = \sum_{i=1}^{g} n\nu\boldsymbol{\omega}_i^g.$$

(Note that the per-site mutation rate, $\nu$, is scaled by the total number of sites in the genome, $n$.)

We also define the rate at which a population in state $\boldsymbol{x}$ transitions to state $\boldsymbol{y}$ by a mutation event (the pairwise mutation rate from $\boldsymbol{x}$ to $\boldsymbol{y}$). There are three conditions that must be satisfied for a population in state $\boldsymbol{x}$ to be allowed to mutate into state $\boldsymbol{y}$: i) the two populations $\boldsymbol{x}$ and $\boldsymbol{y}$ must only differ by exactly one individual in the "source" genotype (the original genotype of the mutating individual) and the "destination" genotype (the new genotype of the mutating individual); ii) $\boldsymbol{x}$ must have one more individual of the source genotype, and $y$ must have one more individual of the destination genotype, and; iii) the source and destination genotypes must be different at exactly one nucleotide position. The pairwise mutation rate from $\boldsymbol{x}$ to $\boldsymbol{y}$ is then

$$\eta(\boldsymbol{x}, \boldsymbol{y}) = \begin{cases} \nu x_G & \text{if conditions i, ii and iii are satisfied} \\ 0 & \text{otherwise,} \end{cases}$$

where $x_G$ is the number of individuals with the source genotype $G$ in population $\boldsymbol{x}$.

Next, we define the rate at which a population in state $\boldsymbol{x}$ transitions to state $\boldsymbol{y}$ by a birth event. Again, there are three conditions that must be satisified for such an event to be possible: i) the two populations $\boldsymbol{x}$ and $\boldsymbol{y}$ must only differ by exactly one individual in the genotype that is giving birth; ii) $\boldsymbol{x}$ must have one fewer individual of the affected genotype than $\boldsymbol{y}$, and; iii) $\boldsymbol{x}$ must have at least one individual of the affected genotype. The rate of transition events due to birth events is:

$$\kappa(\boldsymbol{x}, \boldsymbol{y}) = \begin{cases} r(G)x_G & \text{if conditions i, ii and iii are satisfied} \\ 0 & \text{otherwise.} \end{cases}$$

Finally, we define the rate at which a population in state $\boldsymbol{x}$ transitions to an invalid state (analogous to the absorbing state in our phylogenetic model). In this context, an invalid transition is one that either involves a sampling event, or that involves a birth event that causes the total number of lineages to exceed $c$ (*i.e.,* a birth event that causes a transition out of the subspace $\Omega^g$). This rate is

$$\xi(\boldsymbol{x}) = \begin{cases} \sum_G \phi x_G & \text{if } \sum_G x_G < c \\ \sum_G \left[\phi + r(G)\right] x_G & \text{if } \sum_G x_G = c. \end{cases}$$

The first case accommodates cases where only sampling events cause transitions to invalid states, while the second accomodates cases where both sampling and birth events cause transitions to invalid states.

To derive the transition probabilities, we introduce the quantity $h_{\boldsymbol{y}}(t)$, which represents the probability that the process is in state $\boldsymbol{y}$ at time $t$. We then consider a small interval of time later, $t + \Delta t$, and write down the corresponding Kolmogorov forward equations as:

$$\begin{aligned} h_{\boldsymbol{y}}(t+\Delta t) = & \overbrace{\left[1 - \Lambda(\boldsymbol{y})\Delta t - \Phi(\boldsymbol{y})\Delta t - N(\boldsymbol{y})\Delta t\right] h_{\boldsymbol{y}}(t)}^{\text{no events in time interval}} - \overbrace{\xi(\boldsymbol{y}) h_{\boldsymbol{y}}(t)\Delta t}^{\substack{\text{invalid transition}\\ \text{in time interval}}} \\ & + \underbrace{\sum_{\boldsymbol{x}\in\Omega^g} \eta(\boldsymbol{x}, \boldsymbol{y}) h_{\boldsymbol{x}}(t)\Delta t}_{\text{mutation events into } \boldsymbol{y}} + \underbrace{\sum_{\boldsymbol{x}\in\Omega^g} \kappa(\boldsymbol{x}, \boldsymbol{y}) h_{\boldsymbol{x}}(t)\Delta t}_{\text{birth events into } \boldsymbol{y}} \end{aligned}$$

Taking the limit of $[h_{\boldsymbol{y}}(t+\Delta t) - h_{\boldsymbol{y}}(t)]/\Delta t$ as $\Delta t \to 0$, we arrive at a system of first order ordinary differential equations (one per element of $\Omega^g$):

$$\frac{\mathrm{d}h_{\boldsymbol{y}}(t)}{\mathrm{d}t} = -\left[\Lambda(\boldsymbol{y}) + \Phi(\boldsymbol{y}) + N(\boldsymbol{y}) + \xi(\boldsymbol{y})\right] h_{\boldsymbol{y}}(t) + \sum_{\boldsymbol{x}\in\Omega^g} \left[\eta(\boldsymbol{x}, \boldsymbol{y}) + \kappa(\boldsymbol{x}, \boldsymbol{y})\right] h_{\boldsymbol{x}}(t)$$

We arrange the vector of probabilities $h_{\boldsymbol{y}}(t)$ in a column vector $\boldsymbol{h}(t)$, one per element in $\Omega^g$. We construct a transition-rate matrix $\mathbf{Q} = \{q_{\boldsymbol{xy}}\}$ with element $q_{\boldsymbol{xy}}$ such that

$$q_{\boldsymbol{xy}} = \begin{cases} -\left[\Lambda(\boldsymbol{y}) + \Phi(\boldsymbol{y}) + N(\boldsymbol{y}) + \xi(\boldsymbol{y})\right] & \text{if } \boldsymbol{x} = \boldsymbol{y} \\ \eta(\boldsymbol{x}, \boldsymbol{y}) + \kappa(\boldsymbol{x}, \boldsymbol{y}) & \text{otherwise} \end{cases}$$

This rate matrix allows us to solve the above system of ODEs for an unknown value $\boldsymbol{h}(t+\tau)$, given an initial value $\boldsymbol{h}(t)$:

$$\boldsymbol{h}(t+\tau) = e^{Q\tau}\boldsymbol{h}(t), \tag{S.10}$$

where $e^{Q\tau} = P(\tau)$ is the transition probability matrix. The elements of $P(\tau)$ are transition probabilities between pairs of states, *i.e.,* $p\big(\boldsymbol{y}(t_2) \mid \boldsymbol{x}(t_1)\big)$ corresponds to the element $P_{\boldsymbol{xy}}(t_2 - t_1)$.

*Sampling probability densities.*—Now we turn to computing the probability density of an observed sampling event at time $t$. First, we define the rate at which a population in state $\boldsymbol{x}$ transitions to state $\boldsymbol{y}$ by a sampling event. There are three conditions that must be satisified for such an event to be possible: 1) the two populations $\boldsymbol{x}$ and $\boldsymbol{y}$ must only differ by exactly one individual of the sampled genotype at time $t$; ii) $\boldsymbol{x}$ must have one more individual of the sampled genotype than $\boldsymbol{y}$, and iii) $\boldsymbol{x}$ must have at least one individual of the sampled genotype. The rate of transition events due to sampling events is:

$$\sigma(\boldsymbol{x},\boldsymbol{y}) = \begin{cases} \phi x_G & \text{if conditions i, ii and iii are satisfied} \\ 0 & \text{otherwise.} \end{cases}$$

Next, we derive Kolmogorov forward equations for the sampling probability over an interval from time $t$ to time $t + \Delta t$ (note that, under the assumption that $\Delta t$ is sufficiently small that only one event of any type can occur in the interval, all events other than sampling occur with probability zero):

$$h_{\boldsymbol{y}}(t + \Delta t) = \sigma(\boldsymbol{z},\boldsymbol{y}) h_{\boldsymbol{z}}(t) \Delta t,$$

where $\boldsymbol{z}$ is the state that differs from $\boldsymbol{y}$ by one individual of the genotype sampled at time $t$ (*i.e.*, $\boldsymbol{z} = \boldsymbol{y} + \boldsymbol{x}(t)$). Again, we take the limit of $[h_{\boldsymbol{y}}(t + \Delta t) - h_{\boldsymbol{y}}(t)]/dt$ as $\Delta t \to 0$, resulting in the set of ODEs:

$$\frac{\mathrm{d}h_{\boldsymbol{y}}(t)}{\mathrm{d}t} = \sigma(\boldsymbol{z},\boldsymbol{y}) h_{\boldsymbol{z}}(t).$$

We constuct a sampling-rate matrix, $S = \{s_{\boldsymbol{x},\boldsymbol{y}}\}$ with elements $s_{\boldsymbol{x},\boldsymbol{y}} = \sigma(\boldsymbol{x},\boldsymbol{y})$. The probability density immediately after the sampling event at time $t$ then

$$\boldsymbol{h}_+(t) = S\boldsymbol{h}_-(t),$$

where $+$ and $-$ represent the probability density immediately before and after the sampling event, respectively.

*Integrating out the root age.*—So far, we have described a recursive algorithm for computing the probability of the data—$\boldsymbol{z}$, $\boldsymbol{y}(\boldsymbol{t_e})$, and $T$—given the age of the first bifurcation event, $t_R$ (the root age). However, in practice, the root age is unknown. We therefore want to integrate out the root age. To achieve this, we must specify a prior distribution over possible values of $t_R$, $g(t_R)$, and then integrate the likelihood function over this prior distribution:

$$f(\boldsymbol{z},\boldsymbol{y}(t_e),T \mid M,\lambda_0,\nu,\phi) = \int_{t_R \in \mathbb{R}^+} f(\boldsymbol{z},\boldsymbol{y}(t_e),T \mid M,\lambda_0,\nu,\phi,t_R) g(t_R) \mathrm{d}t_R \tag{S.11}$$

We solve this integral numerically (described in the Implementation subsection).

#### The Biallelic Count Model

So far, we have described a generic model that treats the state of the process as the number of lineages of each genotype. As we noted above, in principle the state space of this model is infinitely large, because there can be a countably infinite number of lineages of each genotype. However, we were able to bound the state space to genotype count vectors where the total number of lineages does not exceed the number of observed samples, $c$, producing a bounded state subspace $\Omega^g$. This bounding is justified because all lineages are ultimately sampled (either stochastically or at the end of the process), so any outcome that involves more than $c$ lineages could never produce the observed count data.

However, even with this bounding, the state space is vast (eq S.7). Given that the SARS-CoV-2 genome is ≈30 kilobases long (so that there are about $120,000$ possible genotypes), a dataset constiting of two samples already involves a state space of size $|\Omega^g| = 7,200,180,001$. Because computing the likelihood function above involves operations with time complexity on the order of the cube of the state-space size (matrix multiplication), we will clearly have to simplify the state space even further.

First, we imagine that a researcher has some *a priori* hypothesis about a specific TE site. In this case, the genotype corresponds to the nucleotide at one site, so there are only 4 genotypes. However, even then, the size of the state space as a function of the number of samples grows quickly:

$$|\Omega^1| = 5$$
$$|\Omega^2| = 15$$
$$|\Omega^5| = 126$$
$$|\Omega^{10}| = 1001$$
$$|\Omega^{50}| = 316251$$
$$|\Omega^{100}| = 4598126$$

Calculations with such a state space are almost certaintly hopeless for realistically sized datasets.

We therefore consider an alternative biallelic model that significantly reduces the size of the state space. In this case, we assume there is a single TE site, with one nucleotide conferring an increased birth rate ("allele 1"), and the remaining nucleotides not affecting the birth rate ("allele 0"). The state is therefore a vector of length 2, with the first value representing the number of lineages with allele 0 and the second value representing the number of lineages with allele 1. This results in a much reduced state space:

$$|\Omega^1| = 3$$
$$|\Omega^2| = 6$$
$$|\Omega^5| = 21$$
$$|\Omega^{10}| = 66$$
$$|\Omega^{50}| = 1326$$
$$|\Omega^{100}| = 5151$$

This simplified state space is in Fig. S3.

##### *Parameters*

The biallelic model is a special case of the full genotype model presented above. The sampling rate $\phi$ remains the same. However, both the birth rates and mutation rates must be modified. The birth rate

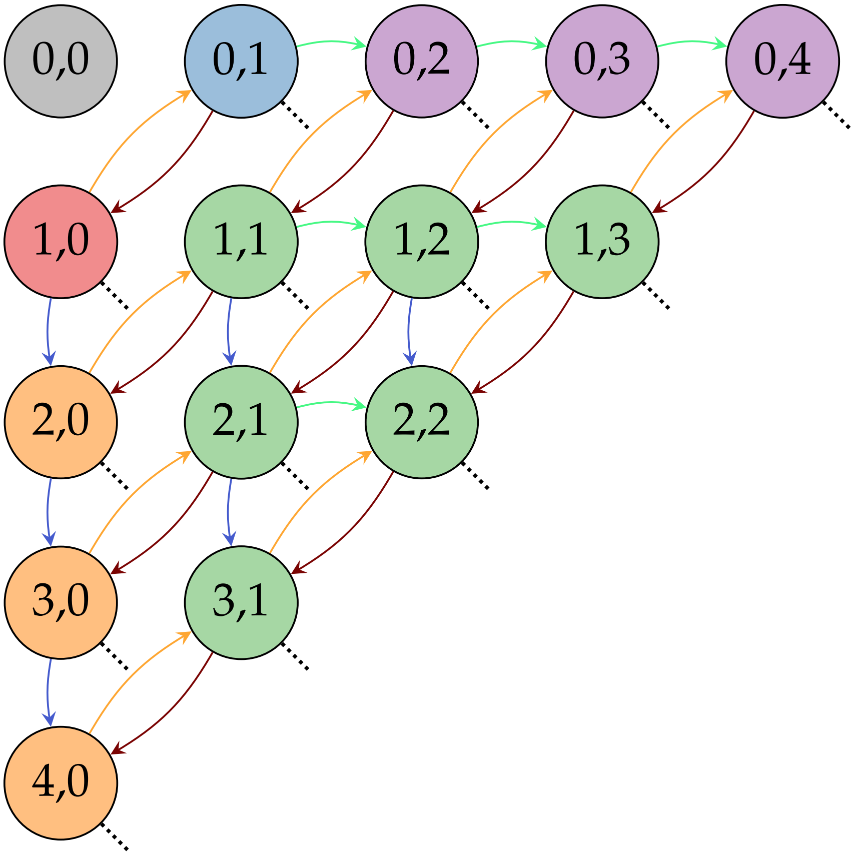


**Figure S3:** Graphical representation of the state space for the biallelic birth-mutation-sampling process with $c = 4$ maximum samples. Nodes represent a state, which consists of the number of lineages with allele 0, $n_0 = i$, and the number of lineages with allele 1, $n_1 = j$. Arrows represent transitions between states. Birth events of lineages with allele 0 (green arrows) occur at rate $i\lambda_0$. Birth events of lineages with allele 1 (blue arrows) occur at rate $j\lambda_1$. Mutation events of lineages with allele 0 to allele 1 (orange arrows) occur at rate $i\nu/3$. Mutation events of lineages with allele 1 to allele 0 (brown arrows) occur at rate $j\nu$. Sampling events, as well as birth events beyond the boundary of $i + j = c$, result in transitions to invalid states (dashed lines). Nodes are color-coded according to which form of ordinary differential equation applies: grey (S.12), red (S.13), blue (S.14), green (S.15), orange (S.16), and purple (S.17).

function $r(G)$ is replaced by two birth rates: $\lambda_0$ for the birth rate of lineages with allele 0, and $\lambda_1$ for the birth rate of lineages with allele 1. Because there are three ways to mutate from one of the non-TE nucleotides to the TE nucleotide, the rate of mutation from 0 to 1 is $3\nu$; mutation events away from the TE nucleotide occur at the usual mutation rate $\nu$.

*Ordinary differential equations and transition probabilities*

In this section we derive ordinary differential equations (ODEs) that describe how the probability of being in state $\boldsymbol{y}(t) = \{n_0 = i, n_1 = j\}$ at time $t$ changes over time. As before, we represent the probability of all states as a column vector $\boldsymbol{h}(t)$, with element $h_{\{i,j\}}(t)$ representing the probability of $\boldsymbol{y}(t) = \{n_0 = i, n_1 = j\}$. While the ODEs for this process are a special case of those we provided in the previous section, we provide explicit formulas for them here because they are more intuitive. Each of these equations applies to a subset of states, corresponding to the relationship between $i$ and $j$ and the boundaries of the state space, as depicted in Fig. S3.

$$\frac{\mathrm{d}h_{\{0,0\}}(t)}{\mathrm{d}t} = 0 \tag{S.12}$$

$$\frac{\mathrm{d}h_{\{1,0\}}(t)}{\mathrm{d}t} = -(\lambda_0 + \frac{\nu}{3} + \phi)h_{\{1,0\}}(t) + \nu h_{\{0,1\}}(t) \tag{S.13}$$

$$\frac{\mathrm{d}h_{\{0,1\}}(t)}{\mathrm{d}t} = -(\lambda_1 + \nu + \phi)h_{\{0,1\}}(t) + \frac{\nu}{3}h_{\{1,0\}}(t) \tag{S.14}$$

$$\frac{\mathrm{d}h_{\{i,j\}}(t)}{\mathrm{d}t} = \left.\begin{array}{l} -(i\lambda_0 + j\lambda_1 + \frac{i\nu}{3} + j\nu + [i+j]\phi)h_{\{i,j\}}(t) \\ +(i-1)\lambda_0 h_{\{i-1,j\}}(t) \\ +(j-1)\lambda_1 h_{\{i,j-1\}}(t) \\ +(i+1)\frac{\nu}{3}h_{\{i+1,j-1\}}(t) \\ +(j+1)\nu h_{\{i-1,j+1\}}(t) \end{array}\right\}\text{for } 1 < i < c,\ 1 < j < c,\ i+j \leq c \tag{S.15}$$

$$\frac{\mathrm{d}h_{\{i,0\}}(t)}{\mathrm{d}t} = \left.\begin{array}{l} -(i\lambda_0 + \frac{i\nu}{3} + i\phi)h_{\{i,0\}}(t) \\ +(i-1)\lambda_0 h_{\{i-1,0\}}(t) \\ +\nu h_{\{i-1,1\}}(t) \end{array}\right\}\text{for } 1 < i \leq c \tag{S.16}$$

$$\frac{\mathrm{d}h_{\{0,j\}}(t)}{\mathrm{d}t} = \left.\begin{array}{l} -(j\lambda_1 + j\nu + j\phi)h_{\{0,j\}}(t) \\ +(j-1)\lambda_1 h_{\{0,j-1\}}(t) \\ +\frac{\nu}{3}h_{\{1,j-1\}}(t) \end{array}\right\}\text{for} 1 < j \leq c \tag{S.17}$$

The corresponding instantaneous rate matrix is very sparse, with non-zero elements:

$$\begin{aligned} Q_{\{i,j\},\{i+1,j\}} &= i\lambda_0 && 0 < i < c,\ 0 \leq j < c-i \\ Q_{\{i,j\},\{i,j+1\}} &= j\lambda_1 && 0 \leq i < c-j,\ 0 < j < c \\ Q_{\{i,j\},\{i-1,j+1\}} &= i\frac{\nu}{3} && 0 < i \leq c,\ 0 \leq j < c-i \\ Q_{\{i,j\},\{i+1,j-1\}} &= j\nu && 0 \leq i < c-i,\ 0 < j \leq c \\ Q_{\{i,j\},\{i,j\}} &= -i\lambda_0 - j\lambda_1 - i\frac{\nu}{3} - j\nu - \big(i+j\big)\phi && 0 < i+j \leq c \end{aligned}$$

For example, consider a model with $c = 2$. We order the possible states $\{0,0\}$, $\{0,1\}$, $\{0,2\}$, $\{1,0\}$,

$\{1,1\}$, and $\{2,0\}$. We assume some initial probability vector $\boldsymbol{h}(t)$. The system of ODEs is:

$$
\begin{aligned}
\frac{\mathrm{d}\boldsymbol{h}(t)}{\mathrm{d}t} &= Q\boldsymbol{h}(t) \\
&= \begin{bmatrix}
0 & 0 & 0 & 0 & 0 & 0 & 0 \\
0 & -(\lambda_1 + \frac{\nu}{3} + \phi) & \lambda_1 & \frac{\nu}{3} & 0 & 0 & \phi \\
0 & 0 & -(2\lambda_1 + 2\frac{\nu}{3} + 2\phi) & 0 & 2\frac{\nu}{3} & 0 & 2\lambda_1 + 2\phi \\
0 & \nu & 0 & -(\lambda_0 + \nu + \phi) & 0 & \lambda_1 & \phi \\
0 & 0 & \nu & 0 & -(\lambda_0 + \lambda_1 + \frac{\nu}{3} + \nu + 2\phi) & \frac{\nu}{3} & \lambda_0 + \lambda_1 + 2\phi \\
0 & 0 & 0 & 0 & 2\nu & -(2\lambda_0 + 2\nu + 2\phi) & 2\lambda_0 + 2\phi \\
0 & 0 & 0 & 0 & 0 & 0 & 0
\end{bmatrix} \times \\
&\quad \begin{bmatrix}
h_{\{0,0\}}(t) \\ h_{\{0,1\}}(t) \\ h_{\{0,2\}}(t) \\ h_{\{1,0\}}(t) \\ h_{\{1,1\}}(t) \\ h_{\{2,0\}}(t)
\end{bmatrix} \\
&= \begin{bmatrix}
0 \\
-(\lambda_1 + \frac{\nu}{3} + \phi)h_{\{0,1\}}(t) + \nu h_{\{1,0\}}(t) \\
-(2\lambda_1 + 2\frac{\nu}{3} + 2\phi) - (\lambda_0 + \nu + \phi) + \lambda_1 h_{\{0,1\}}(t) + \nu h_{\{1,1\}}(t) \\
-(\lambda_0 + \nu + \phi)h_{\{1,0\}}(t) + \frac{\nu}{3} h_{\{1,0\}}(t) \\
-(\lambda_0 + \lambda_1 + \frac{\nu}{3} + \nu + 2\phi)h_{\{1,1\}}(t) + 2\frac{\nu}{3} h_{\{2,0\}}(t) + 2\nu h_{\{0,2\}}(t) \\
-(2\lambda_0 + 2\nu + 2\phi)h_{\{2,0\}}(t) + \lambda_1 h_{\{1,0\}}(()t) + \frac{\nu}{3} h_{\{1,1\}}(t) \\
\phi h_{\{0,1\}}(t) + (2\lambda_1 + 2\phi)h_{\{0,2\}}(t) + \phi h_{\{1,0\}}(t) + (\lambda_0 + \lambda_1 + 2\phi)h_{\{1,1\}}(t) + (2\lambda_0 + 2\phi)h_{\{2,0\}}(t)
\end{bmatrix}
\end{aligned}
$$

which correspond to the set of ODEs described above.

*Sampling events*

As before, we populate a sampling-rate matrix $S(t)$ with non-zero elements:

$$
\begin{aligned}
s_{\{i,j\},\{i-1,j\}}(t) &= i\phi \qquad \text{if } \boldsymbol{x}(t) = \{1,0\} \\
s_{\{i,j\},\{i,j-1\}}(t) &= j\phi \qquad \text{if } \boldsymbol{x}(t) = \{0,1\}
\end{aligned}
$$

#### Implementation

We implemented the biallelic count model in `R` (R Core Team 2022). Here, we provide some critical implementation details of our model, as well as the results of validation tests we performed to ensure that our implementation was correct.

##### *Numerical integration for transition probabilities*

The transition probability, $P(\tau) = e^{Q\tau}$, is fundamental to computing the likelihood under the count model. However, evaluating the matrix exponential for a large instantaneous-rate matrix $Q$ is computationally expensive (the time complexity for most generic algorithms is about $n^3$, where $n$ is the size of the matrix; Moler and Van Loan 2003). Rather than explicitly exponentiating the rate matrix, we solve the system of ODEs, $\frac{\mathrm{d}\boldsymbol{h}(t)}{\mathrm{d}t}$, using a numerical ODE solver. We implemented `c++` versions of our system of ODEs in `R` using the packages `Rcpp` (Eddelbuettel and François 2011) and `RcppEigen` (Bates and Eddelbuettel 2013). We then use the `R` package `deSolve` (Soetaert et al. 2010) to solve this system of ODEs. Specifically, we use the Runga-Kutta-Fehlberg numerical solver (Fehlberg 1969) provided by the function `deSolve::ode`, with absolute and relative error tolerances of $1e^{-6}$ (which proved to be sufficient according to preliminary experiments; results not shown).

##### *Numerical integration for the root age*

The likelihood equation (S.9) assumes the root age, $t_R$, is known. In practice, we do not know the root age, so to compute the likelihood unconditional on the root age, we must assume some prior distribution on the root age and integrate the likelihood over all possible root ages in proportion to this prior, as represented in equation (S.11) (repeated here for convenience):

$$f(\boldsymbol{z}, \boldsymbol{y}(t_e), T \mid M, \lambda_0, \nu, \phi) = \int_{t_R \in \mathbb{R}^+} f(\boldsymbol{z}, \boldsymbol{y}(t_e), T \mid M, \lambda_0, \nu, \phi, t_R) g(t_R) \mathrm{d}t_R$$

However, this integral has no apparent analytical solution. We therefore devised an efficient numerical integration routine to solve this integral.

A naive numerical integration routine would involve recalculating $f(\boldsymbol{z}, \boldsymbol{y}(t_e), T \mid M, \lambda_0, \nu, \phi, t_R)$ for many different values of $t_R$. However, such a naive routine would be extremely inefficient, because many of the computations in $f(\boldsymbol{z}, \boldsymbol{y}(t_e), T \mid M, \lambda_0, \nu, \phi, t_R)$—in particular, transition probabilities and sampling densities—would be redundant.

We devised an efficient numerical integration routine that avoids redundant calculations. Specifically, we specify an ordered set of $n$ candidate values of $t_R$, which we denote $\boldsymbol{t}^C = \{t_1^C, t_2^C, \ldots, t_n^C\}$ (with $t_1^C$ corresponding to the oldest candidate root age). We then compute the likelihood function (S.9) as before, computing sampling and transition probabilities from right (the present) to left (the past), all the way to the oldest candidate root age, $t_1^C$. However, as we move left, we also record values of the conditional likelihood at each candidate root age, $l(\boldsymbol{y}(t_i^C))$. We then compute the integrand of equation (S.11) for each candidate root age using equation (S.8). Finally, we approximate the integral in equation (S.11) using Simpson's rule.

##### *Validation: ODEs*

We validated our system of ODEs by comparing probabilities of states computed using the ODEs against frequency estimates based on Monte Carlo simulations. Specifically, we forward simulated the process beginning at time $t = 0$ with a single lineage in state 0. We then simulated the process

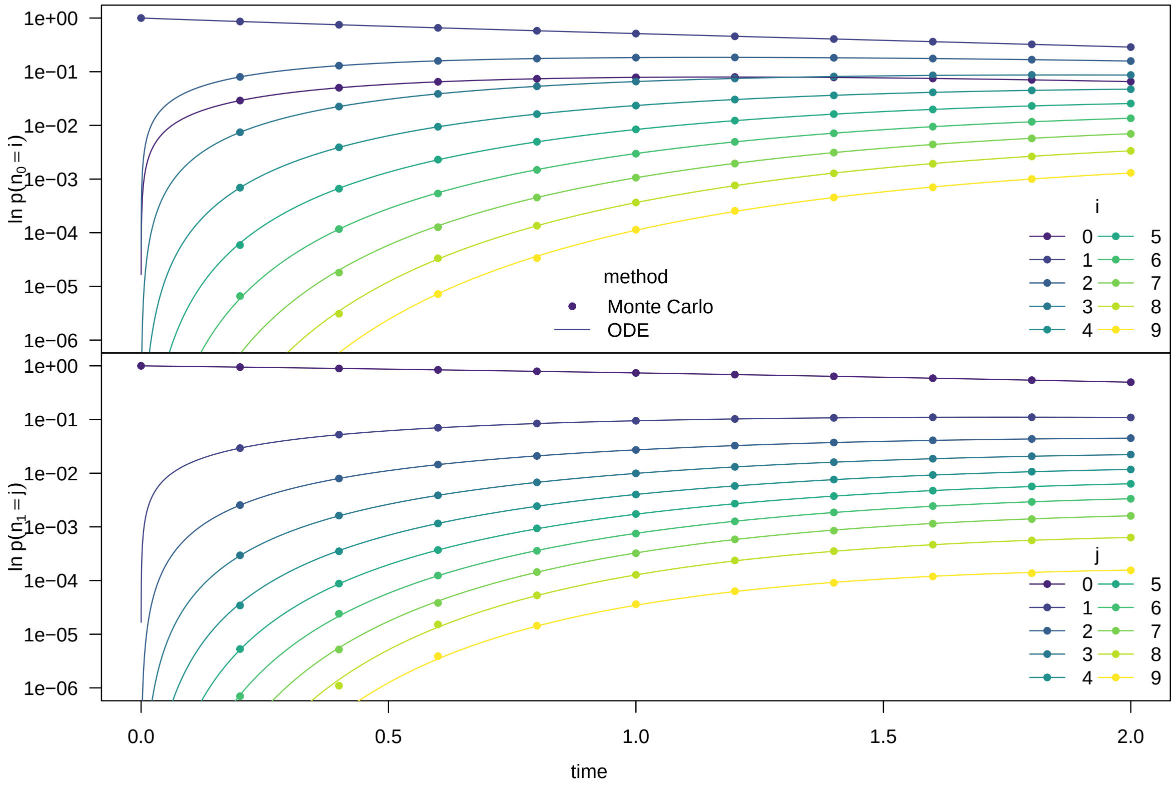


**Figure S4:** The marginal probability over time for the number of lineages in state 0 (top panel) and state 1 (bottom panel). Lines represent to probabilities computed using the ODEs, while dots represent probabilities computed using Monte Carlo simulation.

forward in time, simulating birth events, mutation events, and sampling events for each lineage under the Markov model with rates $\lambda_0 = 0.5$, $\lambda_1 = 1.0$, $\nu = 0.5$, and $\phi = 0.1$. We simulated transitions into the absorbing state if the number of lineages exceeded a critical threshold ($c = 9$), or if the simulation produced a sample. Otherwise, we continued the simulation until it reach the end time, $t_e = 2$. We repeated this procedure 10 million times.

We then computed the marginal frequencies of lineages in state 0 and 1 at each of a set of time points (Fig. S4, dots). The marginal probability of $i$ lineages in state 0 at a particular time is the number of simulations in that state (regardless of the number of lineages in state 1) divided by the total number of simulations (including those that entered the absorbing state by that time).

We then computed the probabilities of each state $\boldsymbol{h}(t)$ for many time points from $t = 0$ to $t = t_e$, according to our ODEs. Specifically, we solved the equation $\boldsymbol{h}(t) = \boldsymbol{h}(0)e^{Qt}$ using a numerical solver, assuming an initial probability vector such that $h_{\{1,0\}}(0) = 1$ and $h_{\{i,j\}}(0) = 0$ for all states $\{i, j\} \neq \{1, 0\}$. This corresponds to beginning with a single lineage in state 0, consistent with our Monte Carlo simulations. We then computed the marginal probabilities of $n_0 = i$ and $n_1 = j$ lineages at time $t$ according to these solutions (Fig. S4, lines).

The marginal probabilities between Monte Carlo simulations and solutions to the ODEs demonstrate nearly perfect agreement (forgiving some Monte Carlo error). This suggests both that our system of ODEs is correct, and that they are correctly implemented.

*Validation: Likelihood*

We validated the likelihood function of the biallelic count model by comparing it to the likelihood of the same data under a biallelic variant of our phylogenetic model that is integrated over all possible trees and node ages, as described eq (S.6). In practice, these integrals are analytically intractable. In each of the scenarios described below, we used multidimensional numerical integration to integrate over the node ages for a given tree topology, and summed these quantities among all possible tree topologies for a given dataset. We checked for consistency between likelihoods for the count model and the (integrated) tree model in three scenarios. We computed likelihoods using the count model and the (integrated) tree model for values of $\lambda_1$ ranging from 0 to 2, assuming $\lambda_0 = 0.5$, $\nu = 0.2$, and $\phi = 0.1$. For simplicity, we assumed the root age was fixed to the true value for both the tree model and the count model.

The likelihoods computed using the count model and (integrated) tree model are essentially identical. This validates the theory and implementation of both our count-based and tree-based likelihood functions.

*Scenario 1.*—For the first scenario, we assumed the process ended at time $t_e = 1$, and the dataset consisted of $\boldsymbol{t} = \{t_1 = 0.5, t_2 = 1\}$ and $\boldsymbol{a} = \{a_1 = 1, a_2 = 0\}$. That is, there was a single serial sample and a single extant sample. In this case, $c = 2$, for which there is just a single tree topology. Likelihoods are identical between the two methods (Fig. S5, top panel).

*Scenario 2.*—For the second scenario, we assumed the process ended at time $t_e = 1.5$, and the dataset consisted of $\boldsymbol{t} = \{t_1 = 0.5, t_2 = 1, t_3 = 1.5\}$ and $\boldsymbol{a} = \{a_1 = 1, a_2 = 0, a_3 = 0\}$. In this case, $c = 3$, for which there are three possible tree topologies. Likelihoods are identical between the two methods (Fig. S5, middle panel).

*Scenario 3.*—For the third scenario, we assumed the process ended at time $t_e = 2$, and the dataset consisted of $\boldsymbol{t} = \{t_1 = 0.5, t_2 = 1, t_3 = 1.5, t_4 = 2\}$ and $\boldsymbol{a} = \{a_1 = 1, a_2 = 0, a_3 = 0, a_4 = 1\}$. In this case, $c = 4$, for which there are 15 posible tree topologies. Likelihoods are identical between the two methods (Fig. S5, bottom panel).

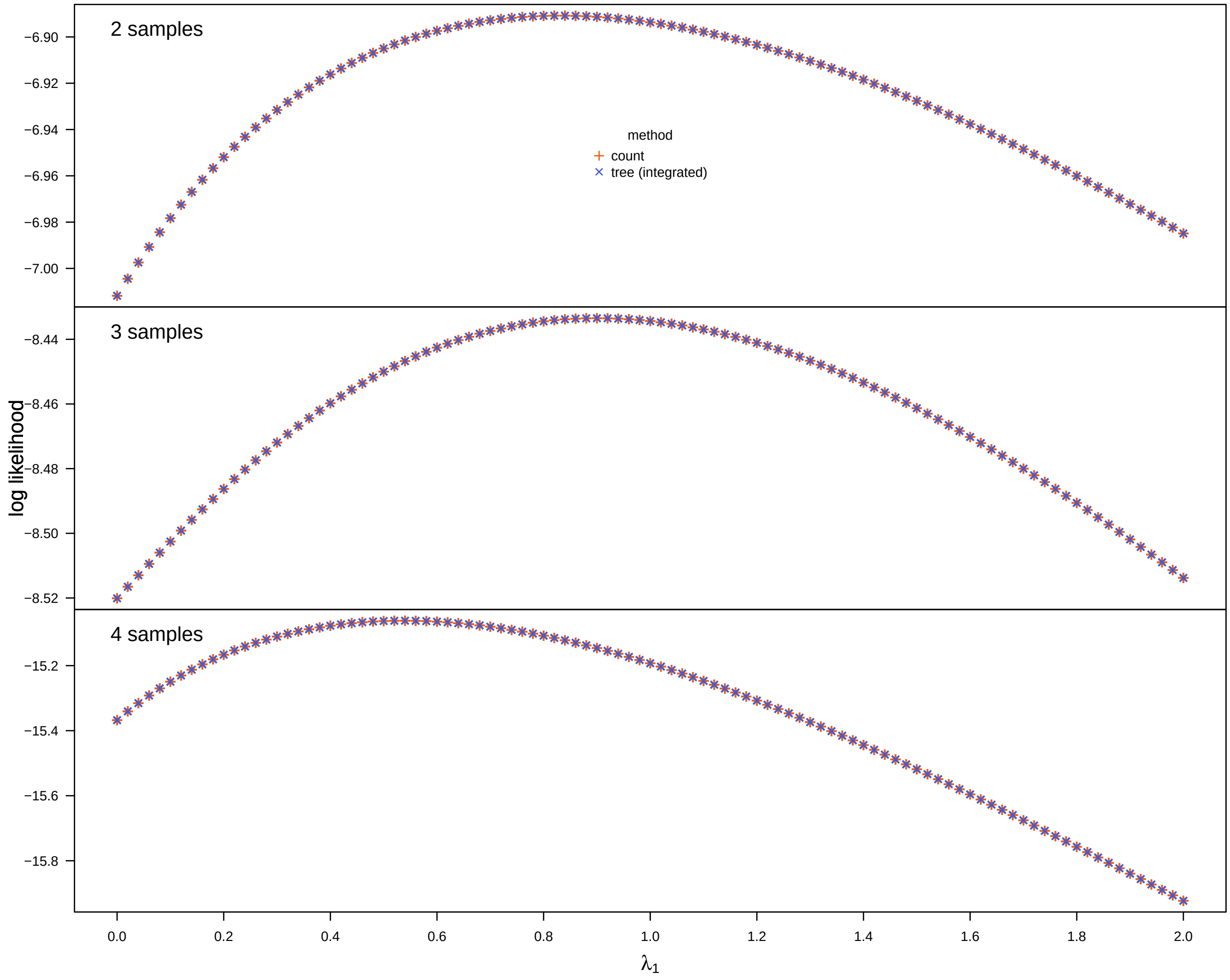


**Figure S5:** Likelihoods ($y$-axis) computed using the biallelic count model compared to those computed using the tree model (integrated over topologies and node ages). We computed likelihood surfaces as a function of $\lambda_1$ ($x$-axis), given fixed values of the remaining parameters ($\lambda_0 = 0.5$, $\nu = 0.2$, and $\phi = 0.1$). We repeated this for three scenarios: one with two samples (one topology, top panel), one with three samples (three topologies, middle panel), and one with four samples (15 topologies, bottom panel). In all cases, the likelihoods are essentially identical.

### S4 Simulation Studies

We performed two simulation studies to understand: 1) the ability to reject neutrality and identify the TE site/allele (Simulation 1), and 2) the relative ability to detect a TE allele between the phylogenetic model and the count model, as a function of time since the origin of the TE allele (Simulation 2). In both cases, we assumed there was a single TE site/allele combination that conferred an increased birth rate. We then used Bayesian inference to compare models and estimate unknown parameters.

#### Simulation 1

To understand the ability to reject a neutral model, and to identify the TE site from among many candidate sites, we simulated trees and nucleotide sequences where one site/allele combination conferred enhanced transmission but the rest were neutral. We simulated trees and genomes under biologically reasonable values of model parameters, with fixed values for the base birth rate, sampling rate, and mutation rate. We varied the value of the TE effect size for allele $j$ at site $i$, $\delta_j(x_{ij})$, and the number of sampled viral genomes, $c$, to assess statistical behavior as a function of the TE effect size and sample size.

##### *Simulations*

We assumed that the sampling rate was $\phi = 1/7$ per day, corresponding to an assumption that a viral infection is sampled after 7 days, on expectation. Based on empirical estimates of the reproductive number for SARS-CoV2, we assumed that was $R_0 = 2.5$ (Kucharski et al. 2020). Normally, the reproductive number is a function of the birth and death rates, *i.e.*, $R_0 = \lambda/\mu$; however, our model does not include death events, which makes it difficult to translate $R_0$ into a biologically meaningful value of $\lambda$. We therefore set $\lambda = R_0 \times \phi$, on the basis that sampling in our model removes a viral infection from circulation, which is similar to the effect of a death event. This translates to a birth rate of $\lambda_0 = 2.5/7 \approx 0.357$ per day. To specify the mutation rate, we used an empirical estimate of the mutation rate of 0.00084 mutations per site per year (following Day et al. 2020, based on Nextstrain [Hadfield et al. 2018] as of May 2020); this corresponds to a mutation rate of $\nu \approx 2.3\times10^{-6}$ per site per day.

We assumed there was a single site in the genome sequence (site $i$ in the sequence $x$) with a single allele that conferred a TE effect (allele $j$). The TE effect size of this site, $\delta_j(x_{ij})$, is the factor by which the birth rate $\lambda_1$ is higher than $\lambda_0$, *i.e.*, $\lambda_1 = \lambda_0 \times \delta_j(x_{ij})$; for simplicity, we refer to this parameter as $\delta$. We arbitrarily fixed the TE site index $i$ to the first position in the genome sequence, and also arbitrarily assumed that nucleotide `A` was the allele that conferred the TE effect. We varied the TE effect size over a range of values, corresponding to $\delta = \{1.25, 1.5, 1.75, 2.5, 3\}$. Finally, we varied the number of samples in the simulated tree, $c = \{100, 200, 400, 800, 1600\}$. For each combination of $\delta$ and $c$, we simulated 1000 datasets according to the procedure described below, for a total of $5 \times 5 \times 1000 = 25,000$ simulated datasets.

We began each simulation with a single lineage. To mimic the gain of a new transmission-enhancing mutation over the course of an outbreak, we drew the initial state for the TE site uniformly at random from among the non-transmission-enhancing nucleotides. We then simulated the mutation and branching process for the TE site forward in time under our Markov model. We simulated this process forward in time until the number of extinct samples plus the number of extant lineages was $c + 1$. We then stopped the simulation, removed the final sample, and chose a new end time uniformly between the stopping time and the time of the previous sampling or speciation event; all of the extinct samples and all of the lineages alive at the new end time were then included in the sampled tree. This procedure guaranteed that the simulation did not end exactly at a sampling or speciation event. We rejected (and reran) simulations where the transmission-enhancing mutation was never gained over the history of the simulation. Finally, we simulated 29,999 neutral sites on the simulated tree under a JC69 model with rate parameter $\nu$, with the initial states drawn from the stationary distribution of the JC69 model (*i.e.*, from a uniform distribution). We then recorded the sequences and tree for each simulation, comprising the total dataset $\mathbf{X}$ and $\Psi$.

*Analysis 1.1: Model Comparison*

Our first set of analyses was designed to assess our ability to identify the site and allele combination that conferred the TE effect. For each simulated dataset, we considered all of the $4^d$ possible non-neutral models (where a given model corresponds to a particular allele at a particular site conferring a TE effect), plus the neutral model (where no site/allele combination confers a TE effect). While we simulated the entire genome of $n$ sites for each lineage, we only consider the $d$ polymorphic sites as candidate TE sites; therefore, the total number of models varies among dataset, ranging from 5 to $\approx$140 (Fig. S6). Recall that the set of TE models is $\mathcal{M} = \{M\}$; we denote the neutral model $M_0$.

In these analyses, we assumed that the mutation rate, $\nu$, the fraction of cases sampled, $\phi$, and the background transmission rate, $\lambda_0$, were known (*i.e.*, fixed to their true values); this reflects the fact that, in empirical applications, researchers will often have good information about the values of these parameters. We then fit each TE model $M$ according to the Bayesian formula:

$$f(M \mid \mathbf{X}, \Psi, \lambda_0, \nu, \phi) = \frac{g(\mathbf{X}^K, \Psi \mid M, \lambda_0, \nu, \phi) h(\mathbf{X}^W, \mathbf{X}^I \mid \Psi, K, \nu) k(M)}{f(\mathbf{X}, \Psi \mid \lambda_0, \nu, \phi)}. \tag{S.18}$$

The first two terms of the numerator are the joint probability of the sequences and phylogeny, as described previously. The last term is the prior on the TE models and parameters, which we describe later. The denominator is the marginal probability of the sequence data and phylogenetic tree, calculated by integrating over all possible models (including the neutral model) and parameters.

The equation (S.18) is a posterior density over a discrete variable (the site index) and a continuous variable (the effect size). However, we are primarily interested in assessing the ability to identify the correct TE site/allele combination, regardless of the specific value of $\delta$. We therefore focus on the integrated posterior model probability, which is the posterior model probability density integrated

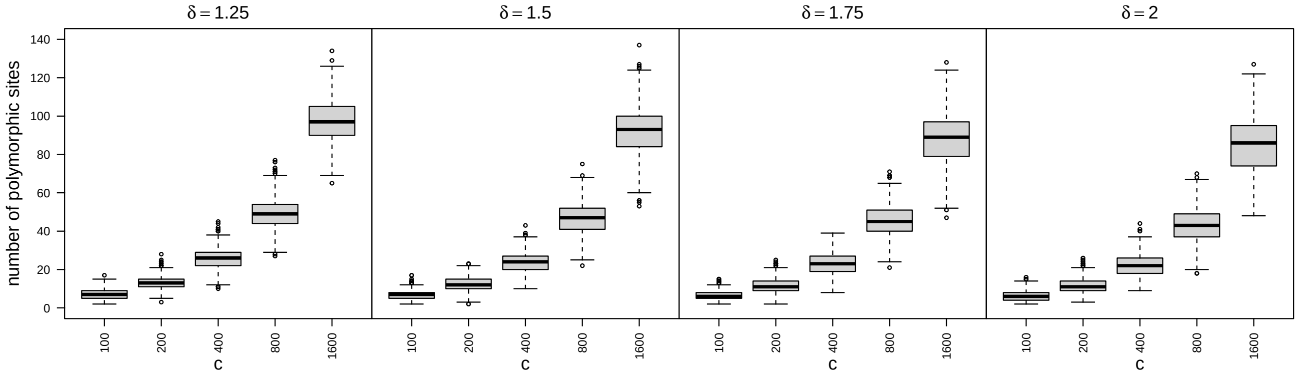


**Figure S6:** The distribution of the number of polymorphic sites in the genome of fixed size $d = 30000$ across simulations ($y$-axis), as a function of the TE effect size ($\delta$, columns) and the number of samples ($c$, $x$-axis).

over all possible values of the TE effect:

$$I(M \mid \mathbf{X}, \Psi, \lambda_0, \nu, \phi) = \int_\delta f(M \mid \mathbf{X}, \Psi, \lambda_0, \nu, \phi)\, \mathrm{d}\delta$$
$$= \frac{\int_\delta g(\mathbf{X}^K, \Psi \mid M, \lambda_0, \nu, \phi) h(\mathbf{X}^W, \mathbf{X}^I \mid \Psi, K, \nu) k(M)\, \mathrm{d}\delta}{f(\mathbf{X}, \Psi \mid \lambda_0, \nu, \phi)} \quad \text{(S.19)}$$

We also fit the neutral model using a Bayesian formula:

$$f(M_0 \mid \mathbf{X}, \Psi, \lambda_0, \nu, \phi) = \frac{q(\Psi \mid \lambda_0, \nu, \phi) h(\mathbf{X}^W, \mathbf{X}^I \mid \Psi, K, \nu) k(M_0)}{f(\mathbf{X}, \Psi \mid \lambda_0, \nu, \phi)}, \quad \text{(S.20)}$$

where $q(\Psi \mid \lambda_0, \nu, \phi) h(\mathbf{X}^W, \mathbf{X}^I \mid \Psi, K, \nu)$, equation (S.5).

Note that the marginal likelihood of both of the above equations is the same, because we include all non-neutral and neutral model. Specifically, the marginal likelihood is:

$$f(\mathbf{X}, \Psi \mid \lambda_0, \nu, \phi) = q(\Psi \mid \lambda_0, \nu, \phi) h(\mathbf{X}^W, \mathbf{X}^I \mid \Psi, K, \nu) k(M_0) +$$
$$\sum_{M \in \mathcal{M}} \int_\delta g(\mathbf{X}^K, \Psi \mid M, \lambda_0, \nu, \phi) h(\mathbf{X}^W, \mathbf{X}^I \mid \Psi, K, \nu) k(M)\, \mathrm{d}\delta \quad \text{(S.21)}$$

*Priors.*—We specify a prior on models such that:

$$k(M) = \begin{cases} 0.5 & \text{if } M = M_0 \\ q(\delta) \times \frac{1}{4^n} & \text{otherwise} \end{cases},$$

which implies that the total probability of all TE models—integrated over the prior density on the effect size, $q(\delta)$—is 0.5. For TE models, we assume:

$$\delta \sim \text{Uniform}(1, 4),$$

*i.e.*, that all possible values of $\delta$ between 1 and 4 are equally likely, such that $q(\delta) = 1/4$.

*Implementation.*—Our goal is to compute the integrated posterior model probability, eq (S.19), which depends primarily on the integral:

$$\int_\delta g(\mathbf{X}^K, \Psi \mid M, \lambda_0, \nu, \phi) h(\mathbf{X}^W, \mathbf{X}^I \mid \Psi, K, \nu) k(M)\, \mathrm{d}\delta$$

for the TE models. We compute this integral using the adaptive numerical integrator function `hcubature` in the `R` package `cubature` (Narasimhan et al. 2022). We then substitute this quantity (and the non-integrated quantity for the neutral model) into equations (S.21), (S.19) and (S.20) to compute the posterior probability of each model.

*Credible sets of models.*—For each analysis, we computed the 95% credible set of models. The 95% credible set is the smallest set of models whose total posterior probability does not exceed 95%. In principle, if the model is true, the credible set should have appropriate frequentist coverage: the true model should be contained in the 95% credible set in 95% of posterior distributions. In order to achieve this interpretation with a discrete set of models, some models have to be included in the credible set stochastically. Imagine we are constructing the 95% credible set and the $k$ highest ranked models have a total posterior probability of $Z < 0.95$, but including the $k+1^{\text{th}}$ model would result in a total posterior probability of greater than 0.95. The first $k$ models are completely contained (they are definitely in the credible set), but the last model may or may not be contained. In these boundary cases, the $k+1^{\text{th}}$ model should be included in the credible set with probability $(0.95 - Z)/P(M_{k+1})$.

For each credible set, we checked whether: 1) the neutral model $M_0$ was in the set; 2) whether the true model $M_{\text{true}}$ was in the set; 3) whether the true model was the model with the highest posterior probability, *i.e.*, it was the maximum *a posteriori* (MAP) model; 4) whether the true model was the only model in the set, *i.e.*, had a posterior probability of about 95%. We plot the frequency of each of these events across simulations as a function of the number of samples and the true effect size.

*Analysis 1.2: Parameter Estimation*

In these analyses, we are interested in posterior estimates of the effect size parameter for the true model, according to the same Bayesian model eq (S.18). For each simulated dataset, we fixed the site/allele combination to the true value, then estimated the posterior distribution of $\delta$ given the model:

$$f(\delta \mid K, \mathbf{X}, \Psi, \lambda_0, \nu, \phi) = \frac{g(\mathbf{X}^K, \Psi \mid M, \lambda_0, \nu, \phi) h(\mathbf{X}^W, \mathbf{X}^I \mid \Psi, K, \nu) q(\delta)}{f(\mathbf{X}, \Psi \mid K, \lambda_0, \nu, \phi)}, \tag{S.22}$$

assuming the same prior density over $\delta$ described previously.

Rather than estimate this parameter using a standard sampling algorithm like Markov chain Monte Carlo, we again used one-dimensional numerical integration functions in the `R` package `cubature` to estimate the mean and variance of the posterior density of $\delta$, as well as the quantile of the true value of $\delta$ within the posterior density.

First, we computed the denominator of eq (S.22):

$$f(\mathbf{X}, \Psi \mid K, \lambda_0, \nu, \phi) = \int_1^4 g(\mathbf{X}^K, \Psi \mid M, \lambda_0, \nu, \phi) h(\mathbf{X}^W, \mathbf{X}^I \mid \Psi, K, \nu) q(\delta) \, \mathrm{d}\delta$$

using one-dimensional numerical integration with the function `hcubature`.

To compute the posterior mean, we solved the integral (again using `hcubature`):

$$\mathbb{E}(\delta; \mathbf{X}^K, \Psi) = \widehat{\delta} = \int_1^4 \delta \left[ \frac{g(\mathbf{X}^K, \Psi \mid M, \lambda_0, \nu, \phi) h(\mathbf{X}^W, \mathbf{X}^I \mid \Psi, K, \nu) q(\delta)}{f(\mathbf{X}, \Psi \mid K, \lambda_0, \nu, \phi)} \right] \mathrm{d}\delta$$

To compute the posterior variance, we solved the integral:

$$\mathrm{Var}(\delta; \mathbf{X}^K, \Psi) = \int_1^4 \left[ \delta - \widehat{\delta} \right]^2 \left[ \frac{g(\mathbf{X}^K, \Psi \mid M, \lambda_0, \nu, \phi) h(\mathbf{X}^W, \mathbf{X}^I \mid \Psi, K, \nu) q(\delta)}{f(\mathbf{X}, \Psi \mid K, \lambda_0, \nu, \phi)} \right] \mathrm{d}\delta$$

Finally, to compute the posterior quantile of the true value, $\delta_{\text{true}}$, we solved the integral:

$$\mathrm{Q}(\delta_{\text{true}}; \mathbf{X}^K, \Psi) = \int_1^{\delta_{\text{true}}} \frac{g(\mathbf{X}^K, \Psi \mid M, \lambda_0, \nu, \phi) h(\mathbf{X}^W, \mathbf{X}^I \mid \Psi, K, \nu) q(\delta)}{f(\mathbf{X}, \Psi \mid K, \lambda_0, \nu, \phi)} \, \mathrm{d}\delta$$

#### Simulation 2

To understand the relative performance of the count method and the tree method in detecting a recently evolved variant of concern, we simulated tree and count data with a single TE allele. As before, we simulated datasets under biologically reasonable values of model parameters, but in this case we only simulated the TE site rather than the entire genome. We varied the value of the TE effect size, and measured the ability to reject a neutral model as a function of the age of the variant.

*Simulations*

We used the same values of $\phi$, $\nu$, and $\lambda_0$ as in Simulation 1. As before, we assumed there was a single site in the genome sequence with a single allele that conferred a TE effect. We varied the TE effect size for the single TE allele over the same range of values, $\delta = \{1.25, 1.5, 1.75, 2.5, 3\}$. Because we analyzed these data under the count model, which is unable to deal with entire genome sequences, we only simulated the TE site (*i.e.*, we ignored the remainder of the genome).

We began each simulation with a single lineage. So that we could precisely control the age of the allele, we assumed the initial lineage began with the TE allele, *i.e.*, our simulation essentially began immediately after the origin of the variant. We then simulated the mutation and branching process for the TE site forward in time under our Markov model. Rather than simulating the process until we collected a fixed number of samples, we simulated for a fixed number of days.

Simulating for a fixed number of days will result in some realizations producing so many samples that the dataset would be too computationally expensive to analyze under the count model. We therefore rejected simulations as soon as they reached 1200 total samples (including serial and terminal samples). However, this type of rejection has the potential to bias our results, because we preferentially retain outcomes that produced a smaller number of samples. To control the impact of this rejection, we adjust the length of the simulation as a function of $\delta$. For a given value of $\delta$, we performed a set of pilot simulations to determine the day at which 5% of the realizations would exceed 1200 samples; we then used this day as the fixed duration for all simulations with that value of $\delta$. The resulting durations for each value of $\delta$ were: 19 days for $\delta = 1.25$, 14 days for $\delta = 1.5$, 12 days for $\delta = 1.75$, and 10 days for $\delta = 1.25$. We also rejected simulations that died (*i.e.*, the last lineage was sampled) before reaching the last day of the simulation.

We simulated 1000 realizations for each value of $\delta$, rejecting and resimulating if the process went extinct or became too large, as described above. For each realization, we recorded both a phylogenetic and count dataset at one day intervals. This resulted in a total of $19 + 14 + 12 + 10 \times 1000 = 55,000$ phylogenetic and count datasets each. We summarized the frequency with which we rejected individual simulations either because they went extinct or became too large (Fig. S7). Note that the while rejection rate because they were too large was $\lesssim$5%, the rate of rejection induced by extinction was $\gtrsim$20%; however, variants that go extinct are not of direct interest because they pose no public health threat, and therefore we are not concerned about any biases induced by this type of rejection.

*Analysis*

We analyzed each day of each dataset under two models: a neutral model, $M_0$, and the true model (where the TE allele was assumed to be known), $M_{\text{true}}$. We assumed the same Bayesian models as in Simulation 1, except that we used the biallelic variant of both the count and phylogenetic models, and we assumed the process began in the TE allele (to match our simulation settings) rather than any of the non-TE alleles. We computed the probability of the counts under a neutral count model by setting $\delta = 1$, *i.e.*, the TE allele conferred no change to the transmission rate. We computed the posterior probability of the (integrated) true model and the neutral model, as described in the previous section.

*Summaries*

For each combination of method (tree or counts), dataset, and day, we checked whether the posterior probability of the true model was at least 95%; in this case, we determined that the method made the correct decision to reject the neutral model. For each combination of method and dataset, we checked whether we rejected the neutral model at all (the true model had a posterior probability of at least 95% for at least one day of the simulation). When a method/dataset combination correctly rejected

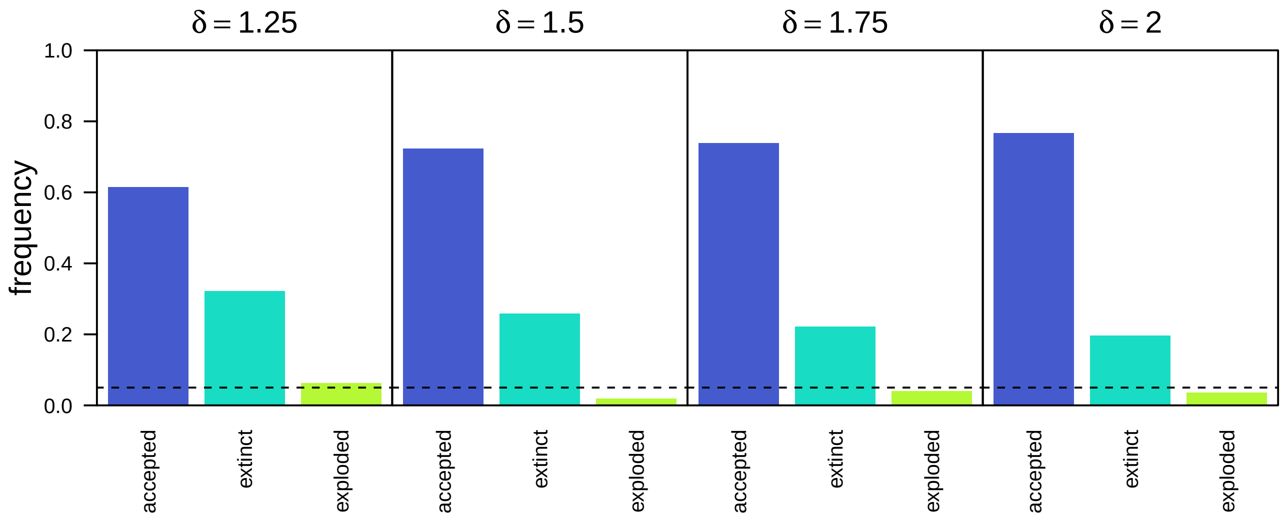


**Figure S7:** Summary of rejection behavior for Simulation 2. We rejected individual simulations if they exceeded 1200 samples ("exploded"), or if the last lineage became a sample before the total duration of the simulation was reached ("extinct"). We calibrated the duration of each simulations so that only ≈5% of the simulations were rejected for being too large.

the neutral model on at least one day, we recorded the earliest day for which we rejected the neutral model. When both count and tree methods rejected the neutral model, we computed the difference in the earliest day for which we rejected the neutral model (the earliest day using the tree method minus the earliest day using the count method).
